# Effectiveness Over Time of a Primary Series of the Original Monovalent COVID-19 Vaccines in Adults in the United States

**DOI:** 10.1101/2024.07.24.24310878

**Authors:** J. Bradley Layton, Patricia C. Lloyd, Lauren S. Peetluk, Yixin Jiao, Djeneba Audrey Djibo, Joann F. Gruber, Jie Deng, Christine Bui, An-Chi Lo, Rachel P. Ogilvie, Ron Parambi, Michael Miller, Jennifer Song, Lisa B. Weatherby, Sylvia Cho, Hui Lee Wong, Tainya C. Clarke, Jessica Rose Hervol, Dóra Illei, Elizabeth J. Bell, Grace Wenya Yang, John D. Seeger, Michael Wernecke, Morgan M. Richey, Richard A. Forshee, Steven A. Anderson, Yoganand Chillarige, Cheryl N. McMahill-Walraven, Kandace L. Amend, Mary S. Anthony, Azadeh Shoaibi

**Affiliations:** RTI Health Solutions, Research Triangle Park, North Carolina, United States; US Food and Drug Administration, Center for Biologics Evaluation and Research, Silver Spring, MD, United States; Optum Epidemiology, Boston, MA, United States; Acumen LLC, Burlingame, CA, United States; CVS Health, Safety Surveillance & Collaboration, Blue Bell, PA, United States; RTI International, Washington, DC, United States; OptumServe, Falls Church, VA, United States

**Keywords:** COVID-19 vaccines, public health surveillance, pharmacoepidemiology

## Abstract

With data from 2 US claims databases (Optum, CVS Health) supplemented with Immunization Information System COVID-19 vaccine records, we evaluated overall and time-specific vaccine effectiveness (VE) of an initial primary series for 3 monovalent COVID-19 vaccines—BNT162b2, mRNA-1273, and JNJ-7836735—in adults (18-64 years). Vaccinated individuals were matched to unvaccinated comparators, and we estimated VE against any medically diagnosed COVID-19 and hospital/emergency department (ED)-diagnosed COVID-19. Additionally, we estimated VE by era of predominant variants, in subgroups, and compared across vaccine brands. The cohorts consisted of 341,097 (Optum) and 1,151,775 (CVS Health) matched pairs for BNT162b2; 201,604 (Optum) and 651,545 (CVS Health) for mRNA-1273; and 49,285 (Optum) and 149,813 (CVS Health) for JNJ-7836735. Summary VE estimates from meta-analysis against hospital/ED-diagnosed COVID-19 were: BNT162b2, 77% (95% CI, 76%-78%); mRNA-1273, 84% (95% CI, 83%-85%), JNJ-7836735 66% (95% CI, 63%-68%). VE estimates were higher for hospital/ED-diagnosed COVID-19 than for medically diagnosed COVID-19, and VE estimates were highest in adults receiving mRNA-1273 for both outcomes. VE was sustained for approximately 7 months for medically-diagnosed and up to 9 months for hospital/ED-diagnosed COVID-19. VE differed by brand and variant era. Additional real-world research is needed as new variants and recommendations for updated vaccines have evolved.

## INTRODUCTION

Vaccines for the prevention of coronavirus disease 2019 (COVID-19) became available in the United States (US) through either emergency use authorization or licensure by the US Food and Drug Administration (FDA). This study evaluated the initial, monovalent primary series of the first 3 authorized vaccines in the US—BNT162b2 (Pfizer-BioNTech’s messenger ribonucleic acid [mRNA] COVID-19 vaccine, Comirnaty®), mRNA-1273 (Moderna’s mRNA COVID-19 vaccine, Spikevax®), and JNJ-7836735 (Janssen Pharmaceutical Company’s adenovirus COVID-19 vaccine). These vaccines demonstrated efficacy in preventing various COVID-19 outcomes, including symptomatic infection,^1–3^ severe COVID-19,^1–4^ and death due to COVID-19^4^ in clinical trials before authorization.

Many early real-world US studies affirmed the real-world effectiveness of COVID-19 vaccines in preventing severe disease,^5–12^ though some suggested that vaccine effectiveness (VE) waned over time in fully vaccinated individuals.^3,7,13–15^ However, some studies demonstrated only minimal or no meaningful decreases in VE estimates over time against more severe COVID-19 or COVID-19-related hospitalization.^5–7,9,16^ In the context of rapidly evolving pandemic conditions, real-world studies on the duration of vaccines’ effectiveness may be challenging. Approaches that compare individuals with different lengths of follow-up may be subject to selection bias and time-related biases (e.g., immortal person-time bias).^17^ Changes in the predominant circulating viral variants further complicate the question of waning effectiveness over time. Specifically, the emergence of Delta and Omicron variants, which emerged several months after vaccination rollout, resulted in large increases in COVID-19 cases among unvaccinated and vaccinated individuals in the US.^18^

FDA continues to monitor the safety and effectiveness of COVID-19 vaccines using real-world evidence through the Biologics Effectiveness and Safety (BEST) Initiative. This study, which is part of the BEST Initiative, evaluated the potential for changing effectiveness of the original complete COVID-19 primary series vaccine over time using rich real-world data and robust methodology.

## MATERIALS AND METHODS

### Data Source and Population

This cohort study of adults aged 18-64 years was conducted using 2 US health insurance claims data sources (Optum pre-adjudicated commercial claims; CVS Health adjudicated commercial claims) supplemented with vaccination records from US Immunization Information Systems (IISs) (sTable 1).^19^ The data sources contain enrollment information, dispensed prescription drug claims, and hospital, physician, and healthcare professional health insurance claims for individuals with commercial insurance from throughout the US. The study period began on 11 December 2020 (the date COVID-19 vaccines were first available in the US) and ended on 15 January 2022 in Optum and 31 March 2022 in CVS Health (end of complete data at the time of analysis). Vaccinated individuals were identified at the first record of a COVID-19 vaccination during the study period (Time 0), and unvaccinated individuals were matched by calendar date, US county of residence, age (5-year increments), sex, immunocompromised status, pregnancy status, history of COVID-19 diagnosis, influenza vaccination, and presence of at least 1 of the conditions identified by CDC as increasing individuals’ risk of severe COVID-19.^20^ The calendar date of the vaccinated individual were set at Time 0 for the matched unvaccinated comparator.

Vaccinated and matched unvaccinated individuals were required to meet the following eligibility criteria on or before Time 0: have continuous medical and pharmacy coverage in the database before Time 0 for at least 365 days prior to Time 0 or back until 11 December 2020 (the day COVID-19 vaccines were initially authorized in the US); be within the indicated/authorized age range of the vaccine; and reside within the catchment area of the combined claims and IIS data. To align vaccinated and unvaccinated individuals, all eligible individuals were required to be free of any of the following conditions in the specified time periods before Time 0, given that these could reduce the probability of vaccination: monoclonal antibody or convalescent plasma treatment (90 days); COVID-19 diagnosis (30 days); fever, nausea/vomiting, rash diagnosis (3 days); hospitalization or emergency department (ED) visit (3 days); or be hospitalized or a long-term care resident (on Time 0).^21^

### Exposure Assessment

Vaccine doses were identified in insurance claims and IIS databases using procedure billing codes for vaccine administration, National Drug Codes for vaccine products, or IIS administration records.^22^ The dose number was inferred from the order of observed doses within an individual’s record—thus continuous enrollment for the entire period since the introduction of vaccines was required. If a vaccine dose of the same brand or a dose record which did not specify the vaccine brand occurred within 3 days following another dose, the second record was considered a duplicate and was excluded; if a dose for a different brand was received within 3 days, the brand of the first dose was considered unknown and both dose records were excluded.

All subsequent COVID-19 vaccines received by an individual after Time 0 were used to evaluate continuing adherence to the primary series (Figure 1). For vaccines with 2-dose primary series, Dose 2 needed to be received no more than 4 days before the recommended interval and no more than 42 days after Dose 1 (sTable 2). A secondary analysis evaluated receipt of only a single dose of the 2-dose mRNA vaccine primary series (Supplemental Methods).

**Figure 1.**
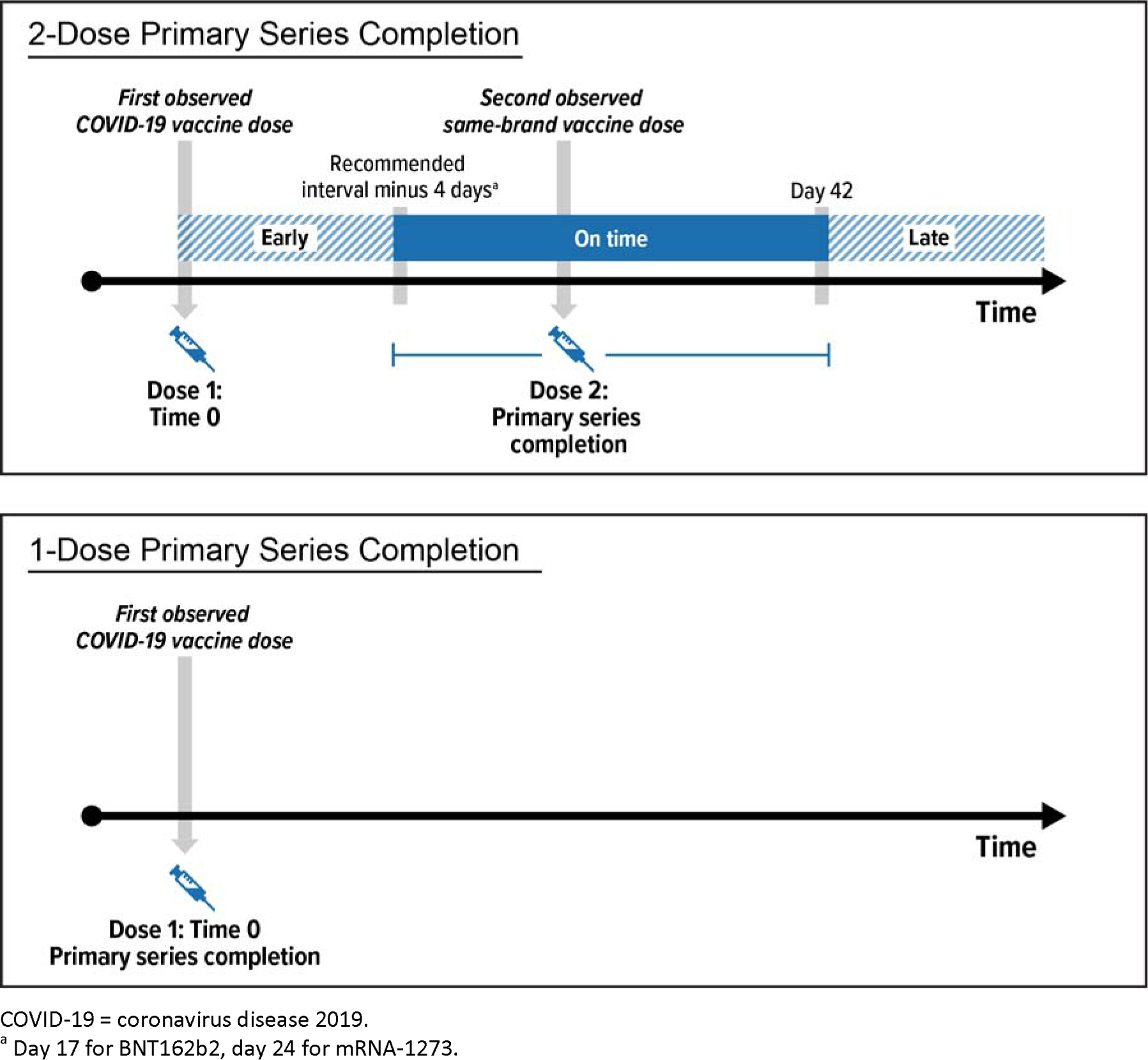
Patterns of Primary Series Completion for 2-Dose and 1-Dose COVID-19 Vaccines.

### Outcome Assessment

Individuals were followed from Time 0 until the occurrence of an outcome of interest, or were censored at the first of the following: last day of the study period; health plan disenrollment; relocation outside of the claims and IIS data catchment area; death; deviation from receiving the primary series (i.e., receipt of Dose 2 too early, failure to receive Dose 2 on time, receiving a booster/additional dose) for the vaccinated, or receipt of any COVID-19 vaccine for the unvaccinated.

Two nested COVID-19 outcomes were evaluated separately: (1) medically diagnosed COVID-19, identified as a recorded COVID-19 diagnosis from hospital, ED, outpatient, or physician encounters; and (2) hospital/ED-diagnosed COVID-19. Recorded COVID-19 diagnosis codes (ICD-10-CM U07.1) were identified in claims in any diagnosis field coding position. For each outcome, the date of the first observed qualifying diagnosis was assigned as the outcome date.

### Covariates

Descriptive characteristics were measured on or before Time 0 (Figure 2) using enrollment, diagnosis, procedure, and pharmacy data.^23^ Covariates included demographics, comorbidities, frailty indicators, healthcare utilization, and conditions potentially increasing risk of severe COVID-19.^20^

**Figure 2.**
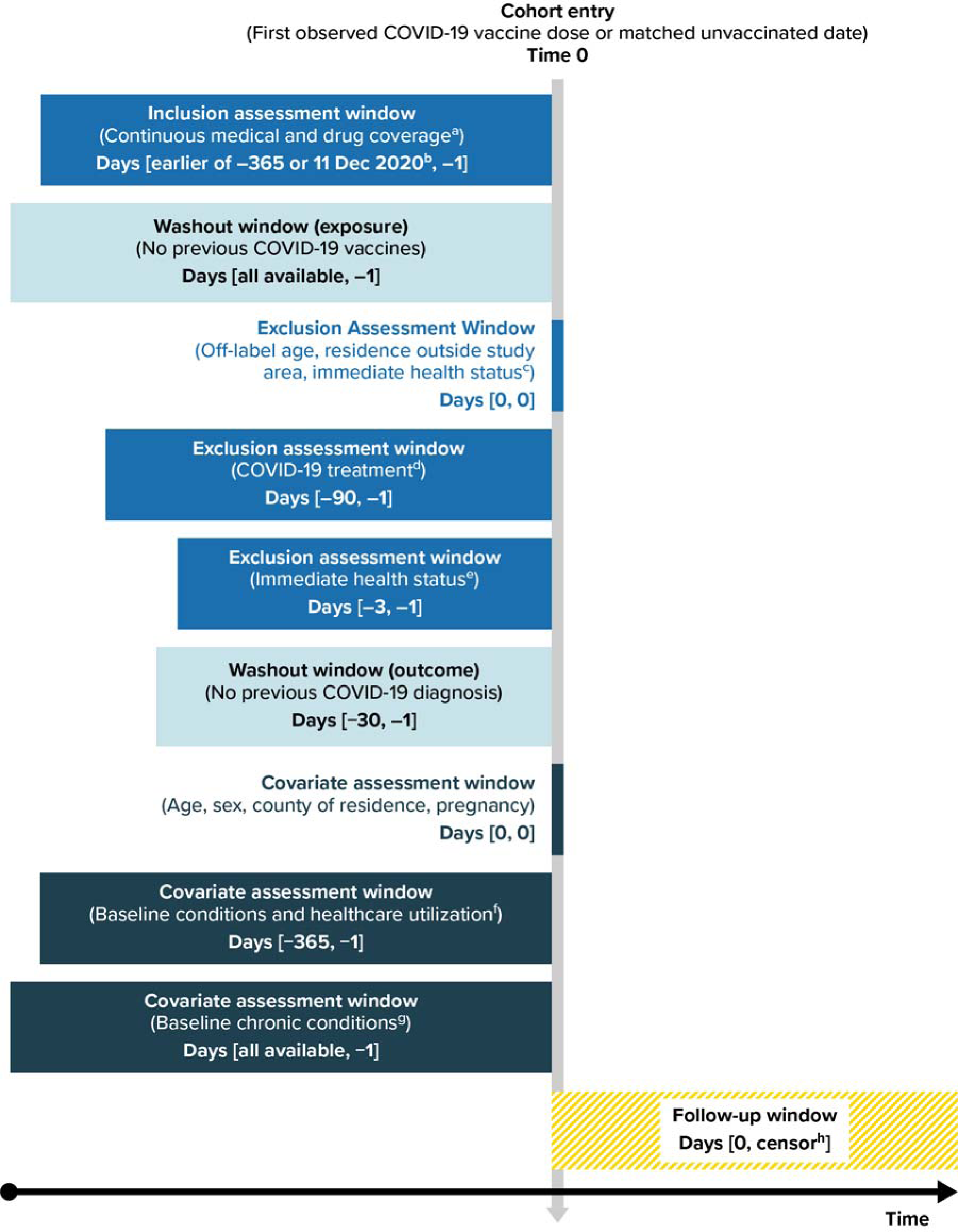
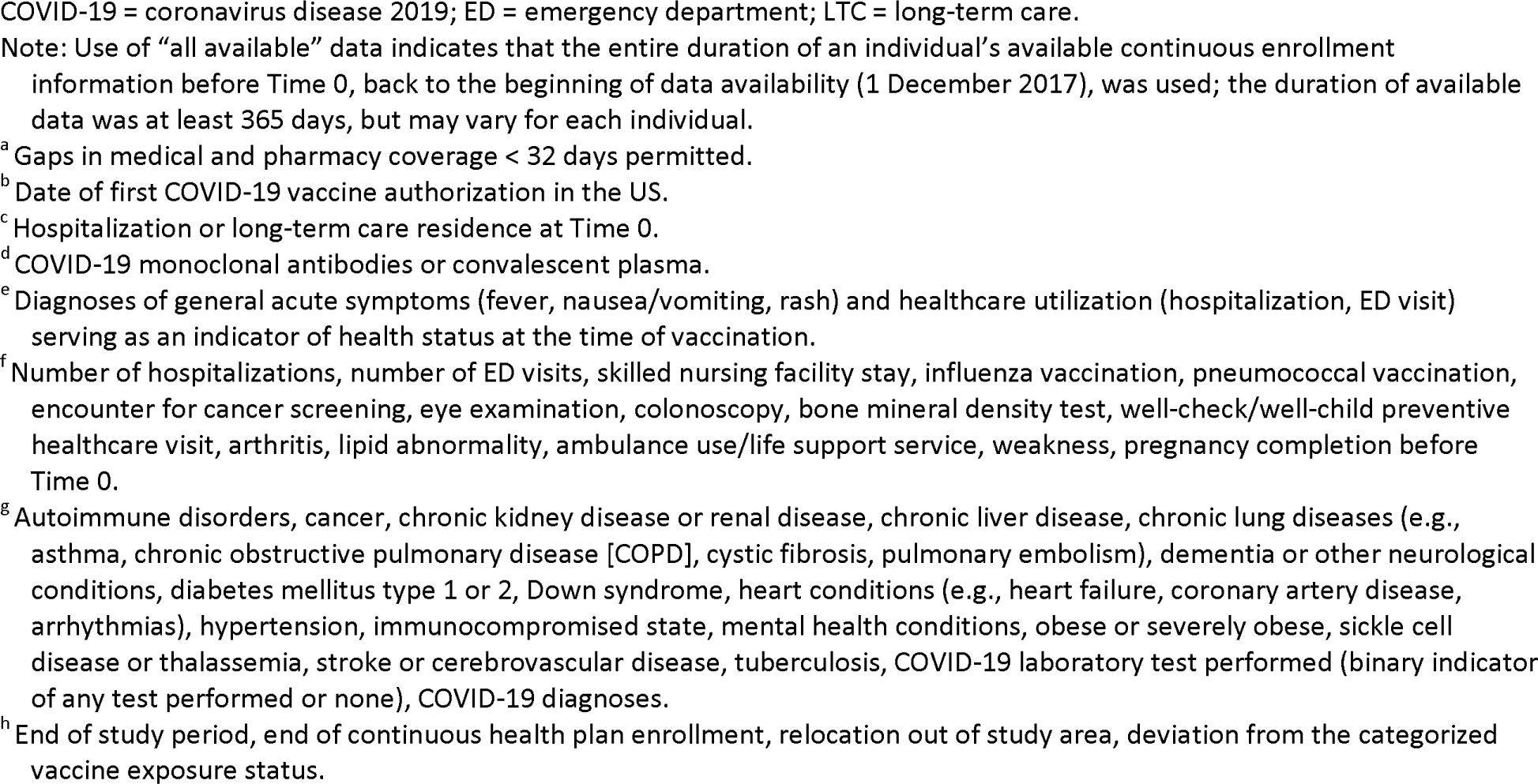
Schematic for Assessing Eligibility, Covariates, and COVID-19 Outcomes Among Vaccinated Individuals and Matched Unvaccinated Comparators.

### Statistical Analysis

Analyses were performed separately by data source and vaccine brand. The distribution of characteristics by vaccination group were described with means, standard deviations (SDs), medians, and first and third quartiles (Q1, Q3) for continuous variables, and counts and proportions for categorical variables. Covariate balance between vaccination groups was evaluated with absolute standardized differences.^24^

In addition to individual matching, propensity score weighting was used to address confounding. Propensity scores were estimated with multivariable logistic regression models including all prespecified covariates and matching factors (sFigure 1). Stabilized inverse probability of treatment (sIPT) weights were estimated from propensity scores with truncation below the 1st percentile and above the 99th percentile of the propensity score distribution.

The cumulative incidence of each COVID-19 outcome was estimated in the sIPT-weighted vaccine exposure groups as 1 minus the Kaplan-Meier estimator.^25^ Cumulative incidence of COVID-19 outcomes in the first 14 days of follow-up was evaluated as a negative control outcome because vaccines are not expected to produce an immune response until 10-14 days after vaccination.^26,27^ Hazard ratios (HRs) for the association of vaccination status with COVID-19 outcomes were estimated using sIPT-weighted Cox proportional hazards models; 95% confidence intervals (CIs) were estimated with robust sandwich variance estimators.^28^ Overall VE was estimated as 1 minus the HR. Time-specific VEs were estimated using daily cumulative incidence estimates in both treatment groups to estimate daily risk ratios (RRs) from which VEs were estimated as 1 minus the RR; time-specific risk differences (RDs) were estimated by subtracting daily cumulative incidence estimates. Nonparametric bootstrapping was used to estimate 95% CIs for time-specific VEs and RDs.

Meta-analyses across data sources were performed with fixed-effects meta-analysis methods^29^ for the overall, subgroup, and variant era–specific VE. Statistical evidence of heterogeneity between data source estimates was evaluated by obtaining log-transformed HR estimates and their standard errors, with *p* values less than 0.05 indicating evidence of statistical heterogeneity.

Subgroup analyses for the overall VE estimation were performed by age, immunocompromised status, and history of COVID-19 diagnosis. Analyses by variant era (pre-Delta era, 11 December 2020 through 31 May 2021; Delta era, 1 June 2021 through 24 December 2021; Omicron era, 25 December 2021 through the end of data availability^30^) were restricted to individuals with Time 0 within the era, and follow-up was censored on the last day of the era. Two sensitivity analyses evaluated the potential selection bias introduced by censoring for receipt of a censoring dose (e.g., individuals in the unvaccinated group receiving any vaccine, or individuals in the vaccinated group receiving Dose 2 too early, Dose 2 of a different brand, or a third dose): (1) delay censoring for 7 days after receipt of a censoring vaccine dose, as there would not be an expected effect during this time; and (2) apply inverse probability of censoring weights.

Quantitative bias analyses (QBA)^31,32^ estimated the impact of potential exposure misclassification because of missing vaccine records by estimating corrected HRs accounting for a range of vaccine exposure sensitivities (Supplemental Methods).

A secondary analysis evaluated the relative VE (RVE) of receiving a complete primary series of different COVID-19 vaccine brands. For each comparison, the study period was restricted to calendar time periods when both vaccine brands in that comparison were authorized. To account for variation in local COVID-19 burden between the 2 exposure groups, vaccinated individuals in both exposure groups were exact-matched 1:1 without replacement on calendar week, age, sex, and US county of residence. RVE was estimated as 1 minus the HR for each pairwise comparison.

Statistical analyses were performed with SAS Version 9.4 (SAS Institute Inc., Cary, NC) and R 4.1.2 (R Foundation for Statistical Computing, Vienna, Austria). This activity was conducted as part of the FDA public health surveillance mandate; FDA did not require Institutional Review Board review. Individual-level consent was not required for this analysis of secondary healthcare data. The study protocol was publicly posted on the BEST Initiative webpage.^23^

## RESULTS

### Descriptive Analyses

In Optum, we identified 612,125 individuals who received an eligible first COVID-19 vaccine dose during the study period. After matching, the analytic cohorts consisted of 341,097 in each group for BNT162b2, 201,604 in each group for mRNA-1273, and 49,285 in each group for JNJ-7836735. In CVS Health, we identified 1,979,109 eligible individuals receiving a first COVID-19 vaccine dose during the study period. After matching, the analytic cohorts consisted of 1,151,775 in each group for BNT162b2, 651,545 in each group for mRNA-1273, and 149,813 in each group for JNJ-7836735. The length of follow-up varied across vaccine brands and outcome analyses, but in Optum, median follow-up ranged from 232 to 248 days for the vaccinated groups, and 88 to 114 days for the unvaccinated; in CVS Health, median follow-up ranged from 244 to 258 days in the vaccinated, and from 102 to 129 days in the unvaccinated.

Selected characteristics of individuals who received a COVID-19 vaccine dose and matched unvaccinated comparators are shown in Table 1 (complete details in Supplemental Information sTables 3-5). Across both data sources, the mean age was approximately 42 to 43 years for each vaccine-specific cohort. The BNT162b2 and mRNA-1273 matched cohorts were slightly more female (51%-52%), but the JNJ-7836735 matched cohort was slightly more male (56%). A very small proportion of the cohorts were pregnant at the time of vaccination (approximately ≤ 1%), and approximately 4% of each cohort were immunocompromised. Vaccine and comparator groups were matched exactly on age, sex, geography, pregnancy, COVID-19 history, and immunocompromised status, thus these characteristics were all perfectly balanced in the matched groups. Additionally, all other measured characteristics (i.e., healthcare utilization, comorbidities, frailty markers) were all well balanced between treatment groups as indicated by absolute standardized differences close to 0 (sTable 3 through sTable 5). The substantial overlap of propensity score distributions for all cohorts indicated a high degree of exchangeability between vaccinated and unvaccinated groups for all measured covariates (sFigure 1).

**Table 1.** Selected Characteristics of Adults Aged 18-64 Years Vaccinated with a COVID-19 Vaccine and Matched Unvaccinated Comparators.

| Characteristic | BNT162b2 |  | mRNA-1273 |  | JNJ-7836735 |  |
| --- | --- | --- | --- | --- | --- | --- |
|  | Vaccinated | Matched, unvaccinated comparators | Vaccinated | Matched, unvaccinated comparators | Vaccinated | Matched, unvaccinated comparators |
| **Optum** | **N = 341,097** | **N = 341,097** | **N = 201,604** | **N = 201,604** | **N = 49,285** | **N = 49,285** |
| *Age, years* |  |  |  |  |  |  |
| Median (Q1, Q3) | 42 (32, 53) | 42 (32, 53) | 44 (33, 54) | 44 (33, 54) | 44 (33, 54) | 44 (33, 54) |
| Mean (SD) | 42.03 (12.95) | 42.02 (12.95) | 43.23 (12.95) | 43.19 (12.97) | 42.91 (12.88) | 42.89 (12.87) |
| *Sex, N (%)* |  |  |  |  |  |  |
| Female | 175,029 (51.31%) | 175,029 (51.31%) | 103,947 (51.56%) | 103,947 (51.56%) | 21,443 (43.51%) | 21,443 (43.51%) |
| Male | 166,068 (48.69%) | 166,068 (48.69%) | 97,657 (48.44%) | 97,657 (48.44%) | 27,842 (56.49%) | 27,842 (56.49%) |
| *Region, N (%)* |  |  |  |  |  |  |
| Midwest | 144,657 (42.41%) | 144,657 (42.41%) | 85,564 (42.44%) | 85,564 (42.44%) | 22,568 (45.79%) | 22,568 (45.79%) |
| Northeast | 40,009 (11.73%) | 40,009 (11.73%) | 26,580 (13.18%) | 26,580 (13.18%) | 5,838 (11.85%) | 5,838 (11.85%) |
| South | 86,610 (25.39%) | 86,610 (25.39%) | 47,138 (23.38%) | 47,138 (23.38%) | 10,032 (20.36%) | 10,032 (20.36%) |
| West | 69,821 (20.47%) | 69,821 (20.47%) | 42,322 (20.99%) | 42,322 (20.99%) | 10,847 (22.01%) | 10,847 (22.01%) |
| Pregnant at Time 0, N (%) | 1,837 (1.05%) | 1,837 (1.05%) | 759 (0.73%) | 759 (0.73%) | 72 (0.34%) | 72 (0.34%) |
| Immunocompromised | 15,135 (4.44%) | 15,135 (4.44%) | 9,000 (4.46%) | 9,000 (4.46%) | 1,865 (3.78%) | 1,865 (3.78%) |
| **CVS Health** | **N = 1,151,775** | **N = 1,151,775** | **N = 651,545** | **N = 651,545** | **N = 149,813** | **N = 149,813** |
| *Age, years* |  |  |  |  |  |  |
| Median (Q1, Q3) | 42 (31, 53) | 42 (31, 53) | 44 (33, 55) | 44 (32, 55) | 44 (32, 54) | 44 (32, 54) |
| Mean (SD) | 41.92 (13.22) | 41.91 (13.23) | 43.37 (13.4) | 43.33 (13.42) | 42.98 (13.31) | 42.97 (13.32) |
| *Sex, N (%)* |  |  |  |  |  |  |
| Female | 615,551 (53.44%) | 615,551 (53.44%) | 338,084 (51.89%) | 338,084 (51.89%) | 67,116 (44.80%) | 67,116 (44.80%) |
| Male | 536,224 (46.56%) | 536,224 (46.56%) | 313,461 (48.11%) | 313,461 (48.11%) | 82,697 (55.20%) | 82,697 (55.20%) |
|  | Vaccinated | Matched, unvaccinated comparators | Vaccinated | Matched, unvaccinated comparators | Vaccinated | Matched, unvaccinated comparators |
| *Region, N (%)* |  |  |  |  |  |  |
| Midwest | 206,280 (17.91%) | 206,280 (17.91%) | 114,493 (17.57%) | 114,493 (17.57%) | 28,431 (18.98%) | 28,431 (18.98%) |
| Northeast | 204,777 (17.78%) | 204,777 (17.78%) | 139,585 (21.42%) | 139,585 (21.42%) | 29,728 (19.84%) | 29,728 (19.84%) |
| South | 251,233 (21.81%) | 251,233 (21.81%) | 113,510 (17.42%) | 113,510 (17.42%) | 26,237 (17.51%) | 26,237 (17.51%) |
| West | 489,485 (42.50%) | 489,485 (42.50%) | 283,957 (43.58%) | 283,957 (43.58%) | 65,417 (43.67%) | 65,417 (43.67%) |
| Pregnant at Time 0, N (%) | 6,611 (0.57%) | 6,611 (0.57%) | 2,557 (0.39%) | 2,557 (0.39%) | 274 (0.18%) | 274 (0.18%) |
| Immunocompromised | 49,753 (4.32%) | 49,753 (4.32%) | 31,211 (4.79%) | 31,211 (4.79%) | 5,561 (3.71%) | 5,561 (3.71%) |

COVID-19 = coronavirus disease 2019; IQR = interquartile range; SD = standard deviation.

### Outcome Analyses

In each cohort, incidence of medically diagnosed COVID-19 was much higher than the incidence of hospital/ED-diagnosed COVID-19. For instance, in Optum, the BNT162b2 vaccinated group had 11,399 medically diagnosed cases and 1,066 hospital/ED-diagnosed cases in nearly 70 million person-days of follow-up; in CVS Health, the BNT162b2 vaccinated group had 40,116 medically diagnosed cases and 4,496 hospital/ED-diagnosed cases in nearly 270 million person-days of follow-up (sTable 6).

### Overall Vaccine Effectiveness

Overall VE estimates are summarized in Figure 3 (complete details in sTable 6). Summary VE estimates from meta-analysis against medically diagnosed COVID-19 for receiving the complete primary series compared with being unvaccinated ranged from 38% to 59% (BNT162b2, 50% [95% CI, 50%-51%]; mRNA-1273, 59% [95% CI, 58%-60%]; JNJ-7836735, 38% [95% CI, 36%-40%]). Meta-analysis VE estimates against hospital/ED-diagnosed COVID-19 were higher, ranging from 66% to 84% (BNT162b2, 77% [95% CI, 76%-78%]; mRNA-1273, 84% [95% CI, 83%-85%]; JNJ-7836735, 66% [95% CI, 63%-68%]). For all outcomes and vaccines, Optum VE estimates were slightly higher than those in CVS Health. Because of the precision of the study estimates, most estimates of overall VE demonstrated evidence of statistical heterogeneity, but estimated magnitudes were similar clinically; therefore, summary VE estimates were reported for all estimates as summaries across both data sources, despite evidence of statistical heterogeneity.

**Figure 3.**
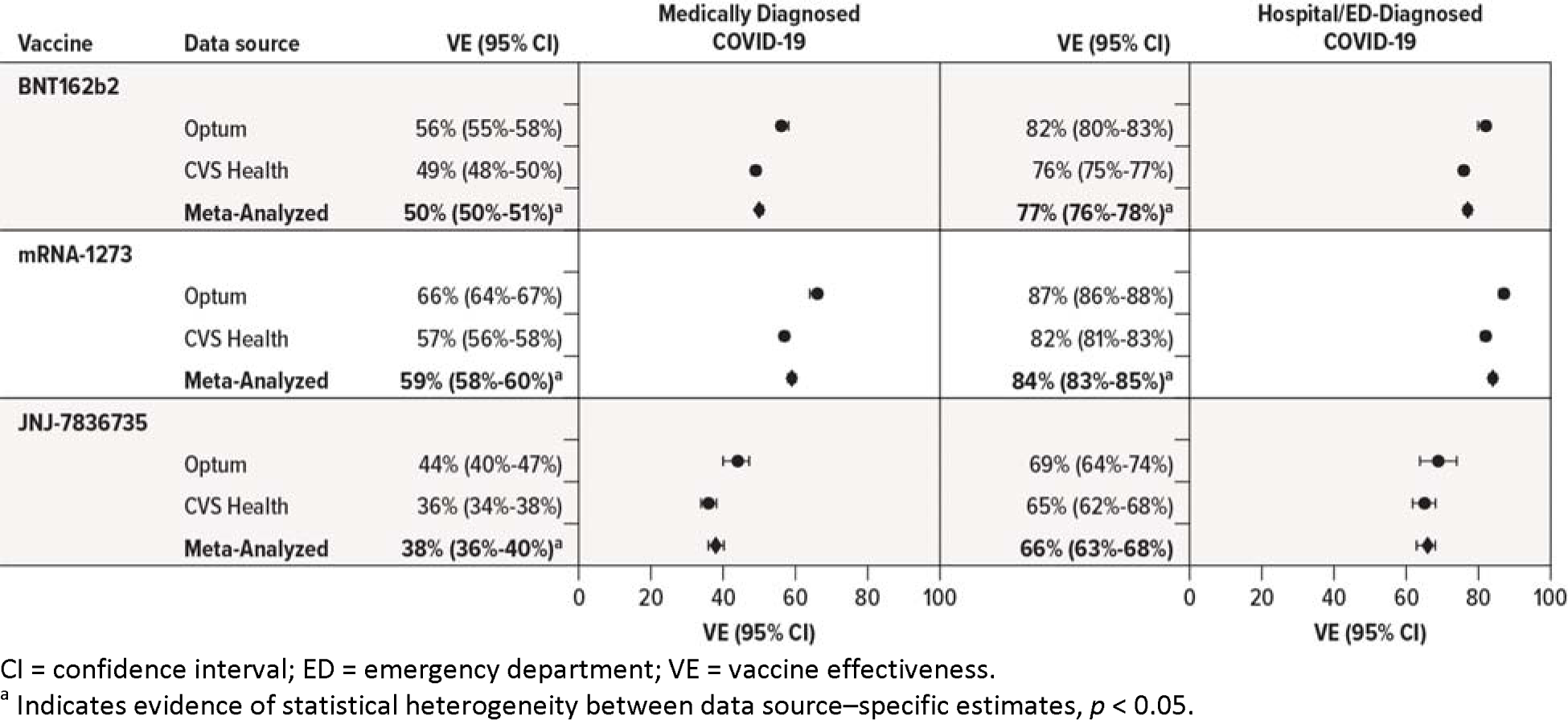
Estimated Effectiveness of Receiving a Complete Primary Series of COVIDD19 Vaccine Compared With Being Unvaccinated in Adults Aged 18-64 Years.

In subgroup analyses, VE estimates for those with a previous COVID-19 diagnosis were generally lower than in those without a previous COVID-19 diagnosis for all brands and both outcomes in both data sources (sFigure 2); however, in the unvaccinated groups, COVID-19 incidence was generally lower among those with a previous COVID-19 diagnosis than among those without a previous diagnosis (sTable 7). Sensitivity analyses were mostly consistent with the primary analyses, but the IPC-weighted analyses resulted in slightly higher VE estimates for the medically diagnosed COVID-19 outcome (sFigure 3). QBA for potentially missing vaccine records suggest the observed overall VE estimates may underestimate the true VE by 2% to 14%, depending on the outcome, analysis, and data source (sTable 8).

### Vaccine Effectiveness Over Time

To evaluate potential changing VE over time for the primary vaccinated versus unvaccinated comparison, the cumulative incidence of COVID-19 outcomes over time by vaccination status and vaccine brand was plotted (sFigure 4). Because of its introduction later in the study period, there was less available follow-up time for the JNJ-7836735 vaccine than the other available vaccines. For each COVID-19 vaccine comparison and outcome, rates were higher in the unvaccinated group throughout follow-up. Generally, differences in outcome rates between the 2 groups were initially small but then widened over time.

**Figure 4.**
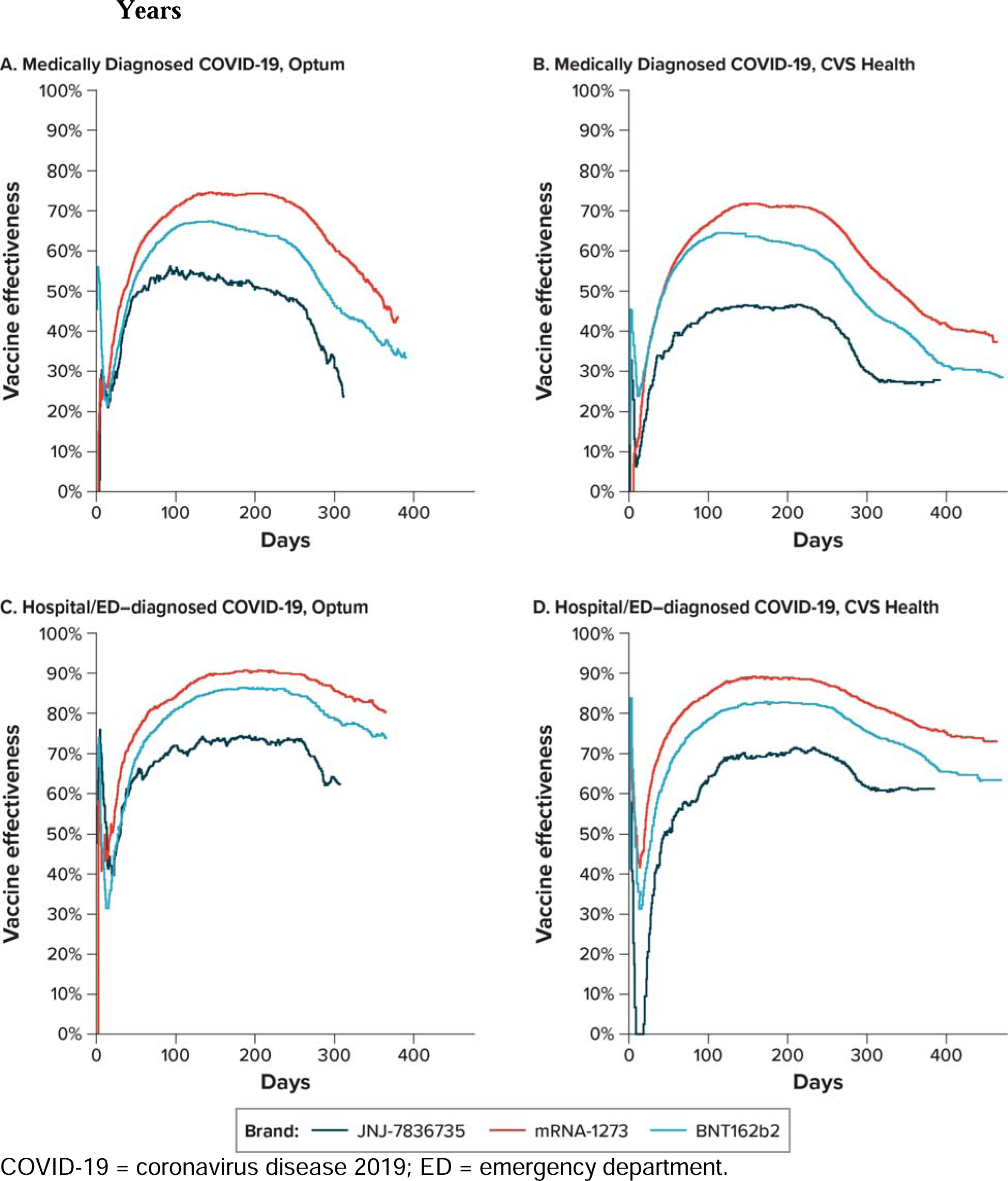
Estimated Effectiveness Over Time of Receiving a Complete Primary Series of COVID-19 Vaccines Compared With Being Unvaccinated in Adults Aged 18-64 Years.

Daily VE estimates throughout follow-up were estimated, and the VE estimates over time were plotted (Figure 4; VE and RD estimates and 95% CIs at specific timepoints are reported in sTable 9). Estimates fluctuated widely during the first week after Time 0 (as the early time periods have the smallest number of events, and estimates were highly subject to change), and negative control outcome analyses of the first 14 days revealed some small differences between vaccinated and unvaccinated groups, mainly in the first few days immediately after vaccination (sFigure 5). Following the initial few days after vaccination, all vaccines followed similar patterns: for medically diagnosed COVID-19, VE estimates increased over time until approximately 90 days and were sustained through approximately day 183, with evidence of waning afterward by day 270; for hospital/ED-diagnosed COVID-19, the pattern was very similar, but the VE estimates were higher throughout, and a smaller degree of waning was observed by day 270 (Figure 4, sTable 9). Despite evidence of waning of the VE estimate, absolute RD estimates showed increasing reductions in numbers of cases through at least day 270 after vaccination (sTable 9).

### Vaccine Effectiveness by Variant Era

VE against both COVID-19 outcomes generally declined between the pre-Delta and Omicron eras (sFigure 6), although there was less available follow-up time during the Omicron era (e.g., only 20 days of follow-up in Optum occurred during the Omicron era), resulting in imprecise estimates. The change in VE between the pre-Delta and Delta eras was greatest for medically diagnosed COVID-19; the meta-analyzed VE of JNJ-7836735 declined by 12 percentage points, followed by BNT162b2 with a 3–percentage point decline. In contrast, the VE of mRNA-1273 increased by 4 percentage points between the pre-Delta and Delta period. Much larger decreases in VE were observed between the Delta and Omicron eras (sFigure 6). For hospital/ED-diagnosed COVID-19, a similar pattern was observed, with only modest changes between the pre-Delta and Delta eras, and more substantial decreases with the Omicron era.

### Relative Vaccine Effectiveness

When estimating the overall RVE for each vaccine brand during time periods when both brands were available, the summary VE estimates for mRNA-1273 demonstrated 16% (95% CI: 15%, 18%) to 19% (95% CI: 14%, 24%) higher effectiveness than BNT162b2 for medically diagnosed and hospital/ED-diagnosed COVID-19, respectively. The relative effectiveness of the 1-dose JNJ-7836735 series was lower than both 2-dose mRNA vaccine brands for both outcomes (sFigure 7).

### Receiving Only One Dose

For the analysis of receiving only a single dose of a 2-dose primary series compared with being unvaccinated, meta-analyzed VE estimates for hospital/ED-diagnosed COVID-19 were 57% (95% CI: 54%, 59%) and 69% (95% CI: 67%, 72%) for single doses of BNT162b2 and mRNA-1273, respectively (sFigure 8). However, these VE estimates for both outcomes were more modest than those seen for the analyses of the complete 2-dose primary series.

## DISCUSSION

In this large, real-world evaluation, receiving a complete primary series of COVID-19 vaccination was effective at preventing medically diagnosed COVID-19 and hospital/ED-diagnosed COVID-19 in adults aged 18 to 64 years, and that effectiveness was sustained for at least 7 months. The observed VE estimates were generally higher for hospital/ED-diagnosed COVID-19 than for any medically diagnosed COVID-19 and were highest in adults receiving the mRNA-1273 vaccine across all analyses. Results from both data sources suggested lower VE in adults receiving the JNJ-7836735 vaccine.

VE was also reduced during the Omicron era across all brands, although sample sizes were too small for the hospital/ED-diagnosed COVID-19 results to be meaningful. When evaluating VE by variant era, VE estimates were lower in the Omicron era than in the Delta or pre-Delta eras for all vaccines and both outcomes. Although waning of VE for the primary series and lower VE in later variant eras was observed, booster doses were recommended and have also been demonstrated to be effective.^33^

Our study utilized 2 US data sources, representing commercially insured adults from jurisdictions across the US with participating IISs who received COVID-19 vaccinations in various healthcare settings and geographic locations. Despite the different data sources, the overall results as well as many of subgroup results were consistent. To summarize the results from these 2 data sources, VE estimates from a meta-analysis are presented. Statistical heterogeneity was observed in many of these results, based on the *I*^2^ statistic, which in some settings would prompt reconsideration of the appropriateness of meta-analysis. However, because our analytical approaches in each data source were identical and the populations similar, we did not find evidence of clinical heterogeneity and thus considered meta-analysis to be a reasonable approach to summarizing, smoothing, and drawing inferences.^34,35^

The eligibility and matching criteria were designed to avoid selection bias by identifying vaccinated and unvaccinated individuals who were eligible for vaccination on each calendar day. Starting follow-up on Time 0 without considering future vaccination behaviors avoided immortal person-time bias by not using future information to define exposure status at baseline.^17^ The daily VE estimates allowed for granular evaluation of changing VE over time.

Initial vaccination rollout focused on higher-risk individuals, but the exact prioritization procedures and groups varied by jurisdiction and changed over time. Because authorizations and recommendations for vaccination changed rapidly during the initial vaccine role out, defining time period- and geography-specific eligibility criteria for the entire study population was impractical; thus, we matched unvaccinated individuals to vaccinated on multiple indicators of eligibility status, including calendar date, immunocompromised status, pregnancy, comorbidities, and age; this matching approximated the rolling eligibility criteria by identifying unvaccinated individuals who were similar to vaccinated individuals on each calendar day in each US county of residence.

Despite the study’s matching on demographic and clinical characteristics and using propensity score weighting, residual confounding may remain. The generally increased VE estimates during the first 14 days of follow-up suggested a potential difference between the exposure groups during a time when vaccines are assumed to have no biologic effect while the body mounts an immune response to a novel antigen. However, other studies^36^ have demonstrated differential COVID-19 testing and diagnoses in the 3-4 days after Time 0, as recently vaccinated individuals may not seek COVID-19 testing, attributing symptoms to vaccine side effects. Additionally, diagnoses of COVID-19 recorded on the same day as vaccination may indicate recording of history of COVID-19,^37^ and these Time 0 events may be more common with vaccine brands administered in healthcare practice settings, rather than pharmacies or mass vaccination centers where concurrent diagnoses cannot be recorded. However, longer-term differences in healthcare-seeking behavior or residual unmeasured confounding cannot be ruled out.

COVID-19 testing capacity and capability have changed rapidly throughout the course of the pandemic, and COVID-19 laboratory testing results were not widely available in the databases utilized for this study, so COVID-19 diagnoses rather than confirmed COVID-19 infections were used as outcomes. COVID-19 severity is not accurately represented in diagnoses,^38^ so we evaluated COVID-19 diagnoses overall in any medically attended setting, and separately in hospital/ED settings as a proxy for severity. However, as the pandemic has progressed, COVID-19–related hospitalizations may be the most relevant public health measure for contemporary surveillance and prevention efforts.^38,39^ We matched vaccination groups on calendar time and evaluated outcomes over calendar time to account for the potentially changing meaning of a COVID-19 diagnosis over time (e.g., care seeking for COVID-19 versus incidental findings).

This real-world study is subject to many limitations common to observational research using existing data sources (e.g., key study elements may be misclassified or missing, the observed VEs may be subject to confounding by unmeasured characteristics, and these results may not be generalizable to other populations). We used health insurance claims supplemented with IIS encounters to identify vaccination status, substantially increasing vaccination capture compared to either data source alone.^19^ However, some vaccines may still be missed, resulting in differential misclassification of exposure. We expect that some truly vaccinated persons may have been classified as unvaccinated due to missing or unidentifiable records, recording errors, or other technical challenges that resulted in incomplete recording of information during the pandemic response. Given this potential for less than 100% sensitivity of our exposure classification records, VE may be underestimated; truly vaccinated individuals, along with their presumably lower risk of COVID-19, would be included in the unvaccinated pool, and thus decrease estimates of VE. To address this potential misclassification, we performed a QBA to determine how our estimates of VE would change based on hypothesized rates of underreporting of vaccination; for medically diagnosed COVID-19, the observed VE may underestimate the true VE by up to 14%. The extent of vaccination completeness and exposure misclassification was estimated using CDC and state-level estimates from the entire population aged less than 65 years, which may not always perfectly align with the study population. Thus, these bias analyses should be interpreted as suggestive of general estimates of direction and magnitude of bias, and not perfectly corrected estimates. The capture of covariates in health insurance diagnosis data may be affected by pandemic-related reductions in healthcare access and utilization, which also may change over the study period.

Receiving a complete primary series of BNT162b2, mRNA-1273, or JNJ-7836735 was associated with reductions in medically diagnosed COVID-19 incidence and hospital/ED-diagnosed COVID-19 in the US adult population, although the observed VE estimates differed across vaccine brands and by variant era. In the rapidly changing dynamics of the COVID-19 pandemic, additional real-world evidence is important for evaluating and understanding changes in VE and impacts of waning immunity on serious COVID-19 outcomes over time.

## Data Availability

The data that support the findings of this study are available from Optum and CVS Health. Restrictions apply to the availability of these data which were authorized for use in this study.

## 1. Financial support

This work was supported by the United States Food and Drug Administration as part of the BEST Initiative.

## 3. The authors thank the following for their contributions

Sarah Harris, MA, of RTI International for project management and support; Dr. Melissa McPheeters, PhD, of RTI International for supervision; Kathryn Matuska, BA, and Wenxuan Zhou, BS, of Acumen LLC for editorial review of the manuscript and interpretation of results; Vivian Wilt, MPH, Julia McIlmail, BS, and Rebecca Warsawski, MPH, of Optum Epidemiology for Project management; Michael Kirksey, MBA, of Optum Enterprise Analytics for data analysis; Dr. Alexandra Stone, PhD, MSc, and Dr. Wafa Tarazi, PhD, MHPA, of Optum Serve Consulting for immunization data integration and quality assurance; Emily Myers, MPH, of Optum Serve Consulting for analytical support; Eli Wolter of Optum Technology for immunization data acquisition and processing; Edward Novak, MS, of Optum Technology for senior technology leadership; Megan Ketchell, BS, of Optum Serve Consulting for project coordination and communications support; Kathryn Federici, MSW, of Optum Serve Consulting for communications lead and legal liaison; Amaka Ume, MPH, of Optum Serve Consulting for analytical support.

## 4. Meeting presentation

Portions of this work have been accepted for a podium presentation at the 2024 ISPE Annual Meeting.

## 5. Disclaimer

This activity was conducted as part of the FDA public health surveillance mandate, and FDA did not require Institutional Review Board review; individual-level consent was not required for this analysis of secondary healthcare data.

## 6. Conflict of interest statement

JBL, CB, DI, MMR, and MSA are employees of RTI International, an independent, nonprofit research institute that performs research on behalf of governmental and commercial clients, including vaccine manufacturers. LP, JD, RO, RP, MM, JS, LBW, GWY, JS, and KA are employees of Optum and own stock in UnitedHealth Group. EJB is an employee of Optum. DAD and CNMW are employees of CVS Health. The other authors have no conflicts to report.

## Abbreviations

CI: confidence interval
COVID-19: coronavirus disease 2019
ED: emergency department
HR: hazard ratio
IIS: Immunization Information Systems
Q1, Q3: first and third quartiles
QBA: quantitative bias analysis
RR: risk ratio
RVE: relative vaccine effectiveness
SD: standard deviation
sIPT: stabilized inverse probability of treatment
US: United States
VE: vaccine effectiveness.

## SUPPLEMENTAL INFORMATION

### Supplemental Methods: Receiving Only 1 Dose of a 2-Dose Primary Series

This secondary analysis evaluated the effectiveness of receiving a single dose of a 2-dose primary series (BNT-162b2 or mRNA-1273), by vaccine brand, compared with being unvaccinated.

The individuals included in this analysis were drawn from the same brand-specific matched analytic cohorts used for the primary analysis (complete vaccine series versus being unvaccinated). Time 0 for both the vaccinated (receipt of Dose 1) and unvaccinated comparator (matched unvaccinated date) groups were the same, and the same 1:1 matching of vaccinated (with an eligible Dose 1 of BNT162b2 or mRNA-1273) and unvaccinated individuals was maintained. However, as this analysis evaluated a different vaccine exposure pattern than the primary analysis (i.e., receiving a single dose of a 2-dose primary series rather than receiving a complete primary series), the censoring criteria for deviation from the vaccine exposure pattern differed for the vaccinated group; vaccinated individuals were censored at the receipt of an additional COVID-19 vaccine dose after Dose 1, as follows:

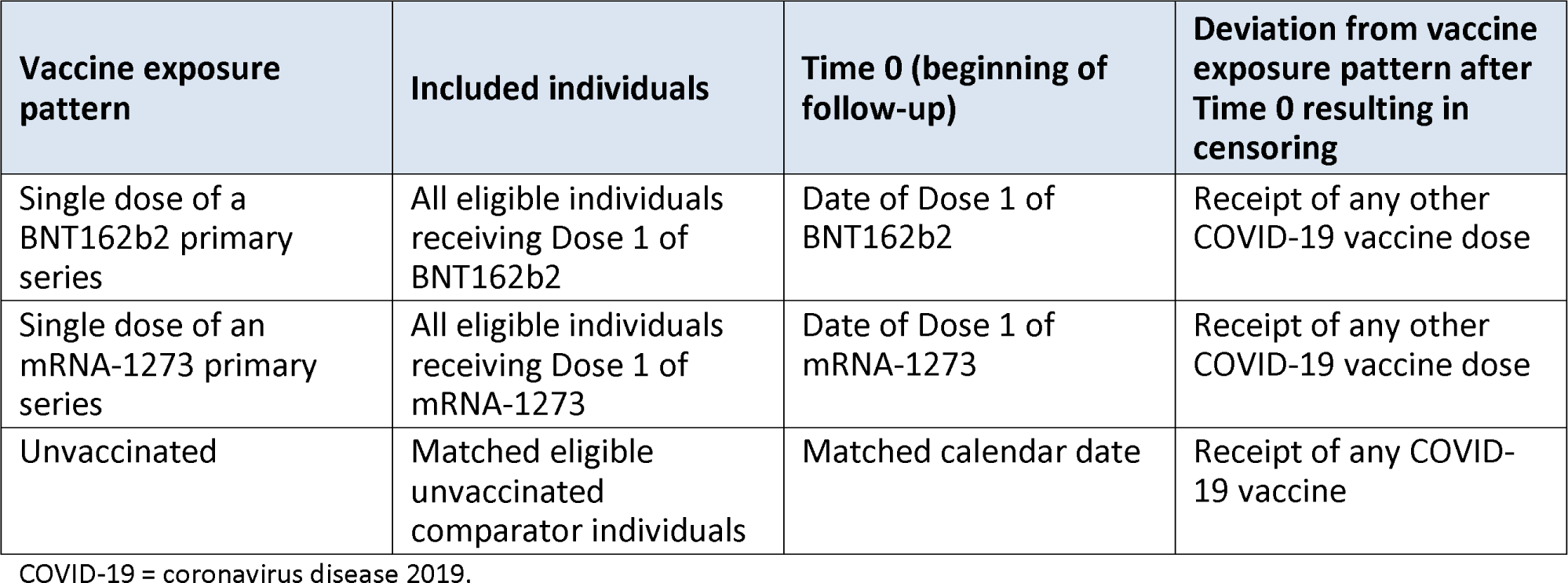

| Vaccine exposure pattern | Included individuals | Time 0 (beginning of follow-up) | Deviation from vaccine exposure pattern after Time 0 resulting in censoring |
| --- | --- | --- | --- |
| Single dose of a BNT162b2 primary series | All eligible individuals receiving Dose 1 of BNT162b2 | Date of Dose 1 of BNT162b2 | Receipt of any other COVID-19 vaccine dose |
| Single dose of an mRNA-1273 primary series | All eligible individuals receiving Dose 1 of mRNA-1273 | Date of Dose 1 of mRNA-1273 | Receipt of any other COVID-19 vaccine dose |
| Unvaccinated | Matched eligible unvaccinated comparator individuals | Matched calendar date | Receipt of any COVID-19 vaccine |

COVID-19 = coronavirus disease 2019.

The same propensity scores and IPT weights used in the primary analysis were used in this analysis. The IPT-weighted cumulative incidence curves of COVID-19 outcomes were plotted by exposure group, and HRs and 95% CIs with the corresponding VE were estimated using IPT-weighted Cox proportional hazards models.

### Supplemental Methods: Quantitative Bias Analysis for Exposure Misclassification

Although vaccinations were identified in both health insurance claims data and IIS COVID-19 vaccine records [1], the possibility for missing vaccine information remains. Quantitative bias analyses [2,3] were performed to estimate the impact of truly vaccinated individuals being misclassified as unvaccinated due to missing vaccine records. No gold standard for vaccine status is available, so estimates of statewide receipt of at least 1 COVID-19 vaccine dose among individuals aged younger than 65 years from CDC, state departments of health, and capture-recapture methods [4-6] were obtained and compared with observed state-level estimates in the study data [1] to estimate maximum and minimum potential sensitivities of vaccine exposure.

Using the minimum and maximum sensitivity estimates from CDC, state departments of health, or capture-recapture methods, 2 “corrected” RR estimates (the primary analyses estimated HRs, but for the purposes of the quantitative bias analyses, RRs and 95% CIs were estimated in the weighted cohorts using a fixed 61-day follow-up time for both outcomes)—a minimum and maximum corrected estimate—were generated for each outcome by reassigning exposure status from unvaccinated to vaccinated based on the sensitivity estimate. A correction factor was then estimated as follows:

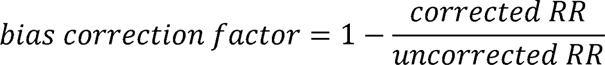

The bias correction factors were then applied to the observed HR estimates from the primary analyses. Specificity of the study’s vaccine assessment was assumed to be 100% (i.e., all observed claims or IIS records were assumed to be true vaccination events, and no truly unvaccinated individuals were misclassified as being vaccinated).

---

**sTable 1.** IIS Jurisdictions and Study Periods Utilized.

| IIS Jurisdiction | Optum Study Period | CVS Health Study Period |
| --- | --- | --- |
| 1 | NA | 11 December 2020 – 31 March 2022 |
| 2 | 11 December 2020 – 15 January 2022 | NA |
| 3 | NA | 11 December 2020 – 31 March 2022 |
| 4 | NA | 11 December 2020 – 31 March 2022 |
| 5 | NA | 11 December 2020 – 31 March 2022 |
| 6 | NA | 11 December 2020 – 31 March 2022 |
| 7 | 11 December 2020 – 15 January 2022 | 11 December 2020 – 31 March 2022 |
| 8 | 11 December 2020 – 15 January 2022 | 11 December 2020 – 31 March 2022 |
| 9 | 11 December 2020 – 15 January 2022 | 11 December 2020 – 31 March 2022 |
| 10 | 11 December 2020 – 15 January 2022 | 11 December 2020 – 31 March 2022 |
| 11 | 11 December 2020 – 15 January 2022 | NA |
| 12 | 11 December 2020 – 15 January 2022 | 11 December 2020 – 31 March 2022 |
| 13 | 11 December 2020 – 15 January 2022 | NA |
| 14 | 11 December 2020 – 15 January 2022 | NA |
| 15 | 11 December 2020 – 15 January 2022 | NA |
| 16 | NA | 11 December 2020 – 31 March 2022 |

IIS = Immunization Information System; NA = not applicable.
Note: Optum used data from 10 IIS jurisdictions from 10 US states; CVS Health used data from 11 IIS jurisdictions from 9 unique US states. Sixteen unique IIS jurisdictions were used in the study.

**sTable 2.** Details of Follow-up for the Complete Primary Vaccination Series Exposure Patterns.

| Vaccine exposure pattern | Included individuals | Time 0 (beginning of follow-up) | Deviation from vaccine exposure pattern after Time 0 resulting in censoring |
| --- | --- | --- | --- |
| BNT162b2 complete primary series | All eligible individuals receiving Dose 1 of BNT162b2 | Date of Dose 1 of BNT162b2 | • Receipt of Dose 2 of BNT162b2 < 17 days after Dose 1<br>• Failure to receive Dose 2 of BNT162b2 by day 42<sup>a</sup><br>• Receipt of any other COVID-19 vaccine brand or unspecified brand<br>• Receipt of a third dose |
| mRNA-1273 complete primary series | All eligible individuals receiving Dose 1 of mRNA-1273 | Date of Dose 1 of mRNA-1273 | • Receipt of Dose 2 of mRNA-1273 < 24 days after Dose 1<br>• Failure to receive Dose 2 of mRNA-1273 by day 42<sup>a</sup><br>• Receipt of any other COVID-19 vaccine brand or unspecified brand<br>• Receipt of a third dose |
| JNJ-7836735 complete primary series | All eligible individuals receiving Dose 1 of JNJ-7836735 | Date of Dose 1 of JNJ-7836735 | • Receipt of any other COVID-19 vaccine dose |
| Unvaccinated | Matched eligible unvaccinated comparator individuals | Matched calendar date | • Receipt of any COVID-19 vaccine dose |

COVID-19 = coronavirus disease 2019.
<sup>a</sup> A 42-day maximum time period was considered for receiving Dose 2 of a 2-dose series.

**sTable 3.** Characteristics of Adults Aged 18-64 Years Vaccinated With BNT162b2 COVID-19 Vaccine and Matched Unvaccinated Comparators.

**A. Optum**
| Characteristic | Individuals vaccinated with BNT162b2<br>N = 341,097 | Matched unvaccinated individuals<br>N = 341,097 | ASD |
| --- | --- | --- | --- |
| **Characteristics assessed at Time 0** |  |  |  |
| *Age, years* |  |  |  |
| Median (Q1, Q3) | 42 (32, 53) | 42 (32, 53) |  |
| Mean (SD) | 42.03 (12.95) | 42.02 (12.95) | 0.00 |
| *Sex, N (%)* |  |  |  |
| Male | 166,068 (48.69%) | 166,068 (48.69%) | 0.00 |
| Female | 175,029 (51.31%) | 175,029 (51.31%) | 0.00 |
| *Region, N (%)* |  |  |  |
| Northeast | 40,009 (11.73%) | 40,009 (11.73%) | 0.00 |
| South | 86,610 (25.39%) | 86,610 (25.39%) | 0.00 |
| Midwest | 144,657 (42.41%) | 144,657 (42.41%) | 0.00 |
| West | 69,821 (20.47%) | 69,821 (20.47%) | 0.00 |
| Pregnant at Time 0, N (%) | 1,837 (0.54%) | 1,837 (0.54%) | 0.00 |
| **Characteristics assessed in the 365 days before Time 0, N (%)** |  |  |  |
| *Hospitalizations* |  |  |  |
| 0 | 231,487 (67.87%) | 231,943 (68.00%) | 0.00 |
| 1 | 60,877 (17.85%) | 60,921 (17.86%) | 0.00 |
| 2+ | 48,733 (14.29%) | 48,233 (14.14%) | 0.00 |
| *Emergency department visits* |  |  |  |
| 0 | 307,322 (90.10%) | 303,855 (89.08%) | 0.03 |
| 1 | 27,521 (8.07%) | 29,610 (8.68%) | 0.02 |
| 2+ | 6,254 (1.83%) | 7,632 (2.24%) | 0.03 |
| Skilled nursing facility stay | 536 (0.16%) | 254 (0.07%) | 0.02 |
| Influenza vaccination | 132,439 (38.83%) | 132,439 (38.83%) | 0.00 |
| Pneumococcal vaccination | 4,732 (1.39%) | 4,316 (1.27%) | 0.01 |
| Encounter for cancer screening | 96,432 (28.27%) | 91,430 (26.80%) | 0.03 |
| Eye examination | 21,650 (6.35%) | 20,043 (5.88%) | 0.02 |
| Colonoscopy | 14,213 (4.17%) | 13,179 (3.86%) | 0.02 |
| Bone mineral density test | 4,011 (1.18%) | 3,684 (1.08%) | 0.01 |
| Well-check/well-child preventive healthcare visit | 146,908 (43.07%) | 139,588 (40.92%) | 0.04 |
| Arthritis | 44,721 (13.11%) | 45,255 (13.27%) | 0.00 |
| Lipid abnormality | 60,872 (17.85%) | 58,489 (17.15%) | 0.02 |
| Ambulance use or life support services | 5,239 (1.54%) | 6,070 (1.78%) | 0.02 |
| Weakness | 6,580 (1.93%) | 6,492 (1.90%) | 0.00 |
| Pregnancy completion before Time 0 | 7,183 (4.10%) | 7,389 (4.22%) | 0.01 |
| **Characteristics assessed using all available data before Time 0, N (%)** |  |  |  |
| Autoimmune disorders | 15,710 (4.61%) | 15,224 (4.46%) | 0.01 |
| Cancer | 20,072 (5.88%) | 19,337 (5.67%) | 0.01 |
| Chronic kidney disease or renal disease | 6,231 (1.83%) | 6,313 (1.85%) | 0.00 |
| Chronic liver disease | 14,482 (4.25%) | 14,576 (4.27%) | 0.00 |
| Chronic lung diseases (e.g., asthma, COPD, cystic fibrosis, pulmonary embolism) | 36,710 (10.76%) | 36,427 (10.68%) | 0.00 |
| Dementia or other neurological conditions | 23,297 (6.83%) | 23,987 (7.03%) | 0.01 |
| Diabetes mellitus, type 1 or 2 | 25,169 (7.38%) | 24,030 (7.04%) | 0.01 |
| Down syndrome | 83 (0.02%) | 61 (0.02%) | 0.00 |
| Heart conditions (e.g., heart failure, coronary artery disease, arrhythmias) | 45,648 (13.38%) | 46,051 (13.50%) | 0.00 |
| Hypertension | 72,219 (21.17%) | 71,842 (21.06%) | 0.00 |
| Immunocompromised state | 15,135 (4.44%) | 15,135 (4.44%) | 0.00 |
| Mental health conditions | 96,696 (28.35%) | 95,899 (28.11%) | 0.01 |
| Obese or severely obese | 71,294 (20.90%) | 70,829 (20.77%) | 0.00 |
| Sickle cell disease or thalassemia | 816 (0.24%) | 904 (0.27%) | 0.01 |
| Stroke or cerebrovascular disease | 5,025 (1.47%) | 5,159 (1.51%) | 0.00 |
| Tuberculosis | 210 (0.06%) | 223 (0.07%) | 0.00 |
| At least 1 COVID-19 laboratory test performed | 127,334 (37.33%) | 115,959 (34.00%) | 0.07 |
| COVID-19 diagnoses occurring outside a hospital or emergency department setting | 18,045 (5.29%) | 17,947 (5.26%) | 0.00 |
| Hospital/ED-diagnosed COVID-19 | 2,810 (0.82%) | 2,777 (0.81%) | 0.00 |

ASD = absolute standardized difference; COVID-19 = coronavirus disease 2019; Q1, Q3 = first and third quartiles; SD = standard deviation.

B. CVS Health
| Characteristic | Individuals vaccinated with BNT162b2<br>N = 1,151,775 | Matched unvaccinated individuals<br>N = 1,151,775 | ASD |
| --- | --- | --- | --- |
| **Characteristics assessed at Time 0** |  |  |  |
| *Age, years* |  |  |  |
| Median (Q1, Q3) | 42 (31, 53) | 42 (31, 53) | 0.00 |
| Mean (SD) | 41.92 (13.22) | 41.91 (13.23) | 0.00 |
| *Sex, N (%)* |  |  |  |
| Male | 536,224 (46.56%) | 536,224 (46.56%) | 0.00 |
| Female | 615,551 (53.44%) | 615,551 (53.44%) | 0.00 |
| *Region, N (%)* |  |  |  |
| Northeast | 204,777 (17.78%) | 204,777 (17.78%) | 0.00 |
| South | 251,233 (21.81%) | 251,233 (21.81%) | 0.00 |
| Midwest | 206,280 (17.91%) | 206,280 (17.91%) | 0.00 |
| West | 489,485 (42.50%) | 489,485 (42.50%) | 0.00 |
| Pregnant at Time 0, N (%) | 6,611 (0.57%) | 6,611 (0.57%) | 0.00 |
| **Characteristics assessed in the 365 days before Time 0, N (%)** |  |  |  |
| *Hospitalizations* |  |  |  |
| 0 | 812,173 (70.51%) | 814,894 (70.75%) | 0.01 |
| 1 | 180,041 (15.63%) | 179,387 (15.57%) | 0.00 |
| 2+ | 159,561 (13.85%) | 157,494 (13.67%) | 0.01 |
| *Emergency department visits* |  |  |  |
| 0 | 1,041,322 (90.41%) | 1,027,640 (89.22%) | 0.04 |
| 1 | 88,687 (7.70%) | 96,968 (8.42%) | 0.03 |
| 2+ | 21,766 (1.89%) | 27,167 (2.36%) | 0.03 |
| Skilled nursing facility stay | 837 (0.07%) | 870 (0.08%) | 0.00 |
| Influenza vaccination | 388,801 (33.76%) | 388,801 (33.76%) | 0.00 |
| Pneumococcal vaccination | 15,854 (1.38%) | 14,639 (1.27%) | 0.01 |
| Encounter for cancer screening | 309,502 (26.87%) | 290,259 (25.20%) | 0.04 |
| Eye examination | 89,086 (7.73%) | 80,665 (7.00%) | 0.03 |
| Colonoscopy | 46,035 (4.00%) | 42,535 (3.69%) | 0.02 |
| Bone mineral density test | 16,067 (1.39%) | 14,520 (1.26%) | 0.01 |
| Well-check/well-child preventive healthcare visit | 455,406 (39.54%) | 428,537 (37.21%) | 0.05 |
| Arthritis | 156,828 (13.62%) | 159,792 (13.87%) | 0.01 |
| Lipid abnormality | 215,402 (18.70%) | 207,851 (18.05%) | 0.02 |
| Ambulance use or life support services | 17,857 (1.55%) | 21,695 (1.88%) | 0.03 |
| Weakness | 21,292 (1.85%) | 21,612 (1.88%) | 0.00 |
| Pregnancy completion before Time 0 | 25,139 (2.18%) | 25,011 (2.17%) | 0.00 |
| **Characteristics Assessed Using all Available Data, N (%)** |  |  |  |
| Autoimmune disorders | 56,025 (4.86%) | 54,760 (4.75%) | 0.01 |
| Cancer | 70,717 (6.14%) | 66,968 (5.81%) | 0.01 |
| Chronic kidney disease or renal disease | 25,567 (2.22%) | 26,629 (2.31%) | 0.01 |
| Chronic liver disease | 56,281 (4.89%) | 56,607 (4.91%) | 0.00 |
| Chronic lung diseases (e.g., asthma, COPD, cystic fibrosis, pulmonary embolism) | 131,033 (11.38%) | 132,083 (11.47%) | 0.00 |
| Dementia or other neurological conditions | 85,608 (7.43%) | 87,745 (7.62%) | 0.01 |
| Diabetes mellitus, type 1 or 2 | 88,515 (7.69%) | 86,908 (7.55%) | 0.01 |
| Down syndrome | 427 (0.04%) | 364 (0.03%) | 0.00 |
| Heart conditions (e.g., heart failure, coronary artery disease, arrhythmias) | 163,592 (14.20%) | 166,283 (14.44%) | 0.01 |
| Hypertension | 243,221 (21.12%) | 243,503 (21.14%) | 0.00 |
| Immunocompromised state | 49,753 (4.32%) | 49,753 (4.32%) | 0.00 |
| Mental health conditions | 323,819 (28.11%) | 326,266 (28.33%) | 0.00 |
| Obese or severely obese | 241,844 (21.00%) | 244,264 (21.21%) | 0.01 |
| Sickle cell disease or thalassemia | 4,042 (0.35%) | 3,904 (0.34%) | 0.00 |
| Stroke or cerebrovascular disease | 20,848 (1.81%) | 22,258 (1.93%) | 0.01 |
| Tuberculosis | 1,145 (0.10%) | 1,098 (0.10%) | 0.00 |
| At least 1 COVID-19 laboratory performed | 527,313 (45.78%) | 484,001 (42.02%) | 0.08 |
| COVID-19 diagnoses occurring outside of a hospital or emergency department | 52,778 (4.58%) | 52,350 (4.55%) | 0.00 |
| Hospitalization or emergency department-diagnosed COVID-19 | 8,983 (0.78%) | 9,161 (0.80%) | 0.00 |

ASD = absolute standardized difference; COVID-19 = coronavirus disease 2019; Q1, Q3 = first and third quartiles; SD = standard deviation.

**sTable 4.** Characteristics of Adults Aged 18-64 Years Vaccinated With an mRNA-1273 COVID-19 Vaccine and Matched Unvaccinated Comparators.

**A. Optum**
| Characteristic | Individuals vaccinated with mRNA-1273<br>N = 201,604 | Matched unvaccinated individuals<br>N = 201,604 | ASD |
| --- | --- | --- | --- |
| **Characteristics assessed at Time 0** |  |  |  |
| *Age, years* |  |  |  |
| Median (Q1, Q3) | 44 (33, 54) | 44 (33, 54) |  |
| Mean (SD) | 43.23 (12.95) | 43.19 (12.97) | 0.00 |
| *Sex, N (%)* |  |  |  |
| Male | 97,657 (48.44%) | 97,657 (48.44%) | 0.00 |
| Female | 103,947 (51.56%) | 103,947 (51.56%) | 0.00 |
| *Region, N (%)* |  |  |  |
| Northeast | 26,580 (13.18%) | 26,580 (13.18%) | 0.00 |
| South | 47,138 (23.38%) | 47,138 (23.38%) | 0.00 |
| Midwest | 85,564 (42.44%) | 85,564 (42.44%) | 0.00 |
| West | 42,322 (20.99%) | 42,322 (20.99%) | 0.00 |
| Pregnant at Time 0, N (%) | 759 (0.38%) | 759 (0.38%) | 0.00 |
| **Characteristics assessed in the 365 days before Time 0, N (%)** |  |  |  |
| *Hospitalizations* |  |  |  |
| 0 | 134,597 (66.76%) | 134,239 (66.59%) | 0.00 |
| 1 | 36,616 (18.16%) | 36,784 (18.25%) | 0.00 |
| 2+ | 30,391 (15.07%) | 30,581 (15.17%) | 0.00 |
| *Emergency department visits* |  |  |  |
| 0 | 181,625 (90.09%) | 179,425 (89.00%) | 0.04 |
| 1 | 16,280 (8.08%) | 17,696 (8.78%) | 0.03 |
| 2+ | 3,699 (1.83%) | 4,483 (2.22%) | 0.03 |
| Skilled nursing facility stay | 210 (0.10%) | 123 (0.06%) | 0.02 |
| Influenza vaccination | 84,003 (41.67%) | 84,003 (41.67%) | 0.00 |
| Pneumococcal vaccination | 3,293 (1.63%) | 2,916 (1.45%) | 0.02 |
| Encounter for cancer screening | 59,499 (29.51%) | 57,384 (28.46%) | 0.02 |
| Eye examination | 13,344 (6.62%) | 12,829 (6.36%) | 0.01 |
| Colonoscopy | 8,905 (4.42%) | 8,366 (4.15%) | 0.01 |
| Bone mineral density test | 2,526 (1.25%) | 2,522 (1.25%) | 0.00 |
| Well-check/well-child preventive healthcare visit | 85,408 (42.36%) | 83,689 (41.51%) | 0.02 |
| Arthritis | 27,688 (13.73%) | 27,843 (13.81%) | 0.00 |
| Lipid abnormality | 40,255 (19.97%) | 38,246 (18.97%) | 0.03 |
| Ambulance use or life support services | 3,211 (1.59%) | 3,669 (1.82%) | 0.02 |
| Weakness | 3,741 (1.86%) | 3,908 (1.94%) | 0.01 |
| Pregnancy completion before Time 0 | 3,593 (3.46%) | 4,090 (3.93%) | 0.03 |
| **Characteristics Assessed Using all Available Data, N (%)** |  |  |  |
| Autoimmune disorders | 9,343 (4.63%) | 9,329 (4.63%) | 0.00 |
| Cancer | 12,495 (6.20%) | 12,215 (6.06%) | 0.01 |
| Chronic kidney disease or renal disease | 4,027 (2.00%) | 3,925 (1.95%) | 0.00 |
| Chronic liver disease | 9,056 (4.49%) | 9,026 (4.48%) | 0.00 |
| Chronic lung diseases (e.g., asthma, COPD, cystic fibrosis, pulmonary embolism) | 23,005 (11.41%) | 22,562 (11.19%) | 0.01 |
| Dementia or other neurological conditions | 14,078 (6.98%) | 14,325 (7.11%) | 0.00 |
| Diabetes mellitus, type 1 or 2 | 17,003 (8.43%) | 16,045 (7.96%) | 0.02 |
| Down syndrome | 50 (0.02%) | 41 (0.02%) | 0.00 |
| Heart conditions (e.g., heart failure, coronary artery disease, arrhythmias) | 28,052 (13.91%) | 28,853 (14.31%) | 0.01 |
| Hypertension | 47,534 (23.58%) | 46,870 (23.25%) | 0.01 |
| Immunocompromised state | 9,000 (4.46%) | 9,000 (4.46%) | 0.00 |
| Mental health conditions | 59,501 (29.51%) | 58,206 (28.87%) | 0.01 |
| Obese or severely obese | 44,965 (22.30%) | 44,041 (21.85%) | 0.01 |
| Sickle cell disease or thalassemia | 433 (0.21%) | 450 (0.22%) | 0.00 |
| Stroke or cerebrovascular disease | 3,236 (1.61%) | 3,501 (1.74%) | 0.01 |
| Tuberculosis | 142 (0.07%) | 109 (0.05%) | 0.01 |
| At least 1 COVID-19 laboratory performed | 72,155 (35.79%) | 65,871 (32.67%) | 0.07 |
| COVID-19 diagnoses occurring outside of a hospital or emergency department | 9,428 (4.68%) | 9,380 (4.65%) | 0.00 |
| Hospitalization or emergency department-diagnosed COVID-19 | 1,476 (0.73%) | 1,445 (0.72%) | 0.00 |

ASD = absolute standardized difference; COVID-19 = coronavirus disease 2019; Q1, Q3 = first and third quartiles; SD = standard deviation.

B. CVS Health
| Characteristic | Individuals vaccinated with mRNA-1273<br>N = 651,545 | Matched unvaccinated individuals<br>N = 651,545 | ASD |
| --- | --- | --- | --- |
| **Characteristics assessed at Time 0** |  |  |  |
| *Age, years* |  |  |  |
| Median (Q1, Q3) | 44 (33, 55) | 44 (32, 55) | 0.00 |
| Mean (SD) | 43.37 (13.4) | 43.33 (13.42) | 0.00 |
| *Sex, N (%)* |  |  |  |
| Male | 313,461 (48.11%) | 313,461 (48.11%) | 0.00 |
| Female | 338,084 (51.89%) | 338,084 (51.89%) | 0.00 |
| *Region, N (%)* |  |  |  |
| Northeast | 139,585 (21.42%) | 139,585 (21.42%) | 0.00 |
| South | 113,510 (17.42%) | 113,510 (17.42%) | 0.00 |
| Midwest | 114,493 (17.57%) | 114,493 (17.57%) | 0.00 |
| West | 283,957 (43.58%) | 283,957 (43.58%) | 0.00 |
| Pregnant at Time 0, N (%) | 2,557 (0.39%) | 2,557 (0.39%) | 0.00 |
| **Characteristics in the 365 Days Before Time 0, N (%)** |  |  |  |
| *Hospitalizations* |  |  |  |
| 0 | 456,478 (70.06%) | 453,734 (69.64%) | 0.01 |
| 1 | 103,083 (15.82%) | 103,735 (15.92%) | 0.00 |
| 2+ | 91,984 (14.12%) | 94,076 (14.44%) | 0.01 |
| *Emergency department visits* |  |  |  |
| 0 | 587,420 (90.16%) | 580,072 (89.03%) | 0.04 |
| 1 | 51,337 (7.88%) | 55,763 (8.56%) | 0.02 |
| 2+ | 12,788 (1.96%) | 15,710 (2.41%) | 0.03 |
| Skilled nursing facility stay | 567 (0.09%) | 550 (0.08%) | 0.00 |
| Influenza vaccination | 239,416 (36.75%) | 239,416 (36.75%) | 0.00 |
| Pneumococcal vaccination | 10,819 (1.66%) | 9,675 (1.48%) | 0.01 |
| Encounter for cancer screening | 182,176 (27.96%) | 174,072 (26.72%) | 0.03 |
| Eye examination | 52,718 (8.09%) | 48,741 (7.48%) | 0.02 |
| Colonoscopy | 28,306 (4.34%) | 26,338 (4.04%) | 0.02 |
| Bone mineral density test | 10,114 (1.55%) | 9,424 (1.45%) | 0.01 |
| Well-check/well-child preventive healthcare visit | 251,488 (38.60%) | 241,716 (37.10%) | 0.03 |
| Arthritis | 95,284 (14.62%) | 95,699 (14.69%) | 0.00 |
| Lipid abnormality | 140,791 (21.61%) | 134,374 (20.62%) | 0.02 |
| Ambulance use or life support services | 11,600 (1.78%) | 12,988 (1.99%) | 0.02 |
| Weakness | 12,206 (1.87%) | 12,208 (1.87%) | 0.00 |
| Pregnancy completion before Time 0 | 11,057 (1.70%) | 12,237 (1.88%) | 0.01 |
| **Characteristics Assessed Using all Available Data, N (%)** |  |  |  |
| Autoimmune disorders | 34,439 (5.29%) | 33,671 (5.17%) | 0.01 |
| Cancer | 43,865 (6.73%) | 42,441 (6.51%) | 0.01 |

| **Characteristic** | **Individuals vaccinated with mRNA-1273**<br>**N = 651,545** | **Matched unvaccinated individuals**<br>**N = 651,545** | **ASD** |
| --- | --- | --- | --- |
| Chronic kidney disease or renal disease | 17,312 (2.66%) | 17,201 (2.64%) | 0.00 |
| Chronic liver disease | 35,180 (5.40%) | 35,288 (5.42%) | 0.00 |
| Chronic lung diseases (e.g., asthma, COPD, cystic fibrosis, pulmonary embolism) | 81,057 (12.44%) | 80,729 (12.39%) | 0.00 |
| Dementia or other neurological conditions | 51,559 (7.91%) | 52,066 (7.99%) | 0.00 |
| Diabetes mellitus, type 1 or 2 | 59,510 (9.13%) | 56,297 (8.64%) | 0.02 |
| Down syndrome | 313 (0.05%) | 212 (0.03%) | 0.01 |
| Heart conditions (e.g., heart failure, coronary artery disease, arrhythmias) | 100,214 (15.38%) | 101,431 (15.57%) | 0.01 |
| Hypertension | 158,070 (24.26%) | 156,296 (23.99%) | 0.01 |
| Immunocompromised state | 31,211 (4.79%) | 31,211 (4.79%) | 0.00 |
| Mental health conditions | 193,580 (29.71%) | 191,353 (29.37%) | 0.01 |
| Obese or severely obese | 151,991 (23.33%) | 149,445 (22.94%) | 0.01 |
| Sickle cell disease or thalassemia | 2,011 (0.31%) | 2,055 (0.32%) | 0.00 |
| Stroke or cerebrovascular disease | 14,143 (2.17%) | 14,716 (2.26%) | 0.01 |
| Tuberculosis | 575 (0.09%) | 614 (0.09%) | 0.00 |
| At least 1 COVID-19 laboratory performed | 290,717 (44.62%) | 266,802 (40.95%) | 0.07 |
| COVID-19 diagnoses occurring outside of a hospital or emergency department | 27,212 (4.18%) | 27,004 (4.14%) | 0.00 |
| Hospitalization or emergency department-diagnosed COVID-19 | 4,739 (0.73%) | 4,899 (0.75%) | 0.00 |

ASD = absolute standardized difference; COVID-19 = coronavirus disease 2019; Q1, Q3 = first and third quartiles; SD = standard deviation.

**sTable 5.** Characteristics of Adults Aged 18-64 Years Vaccinated With JNJ-7836735 COVID-19 Vaccine and Matched Unvaccinated Comparators.

**A. Optum**
| Characteristic | Individuals vaccinated with JNJ-7836735<br>N = 49,285 | Matched unvaccinated individuals<br>N = 49,285 | ASD |
| --- | --- | --- | --- |
| **Characteristics assessed at Time 0** |  |  |  |
| *Age, years* |  |  |  |
| Median (Q1, Q3) | 44 (33, 54) | 44 (33, 54) |  |
| Mean (SD) | 42.91 (12.88) | 42.89 (12.87) | 0.00 |
| *Sex, N (%)* |  |  |  |
| Male | 27,842 (56.49%) | 27,842 (56.49%) | 0.00 |
| Female | 21,443 (43.51%) | 21,443 (43.51%) | 0.00 |
| *Region, N (%)* |  |  |  |
| Northeast | 5,838 (11.85%) | 5,838 (11.85%) | 0.00 |
| South | 10,032 (20.36%) | 10,032 (20.36%) | 0.00 |
| Midwest | 22,568 (45.79%) | 22,568 (45.79%) | 0.00 |
| West | 10,847 (22.01%) | 10,847 (22.01%) | 0.00 |
| Pregnant at Time 0, N (%) | 72 (0.15%) | 72 (0.15%) | 0.00 |
| **Characteristics in the 365 Days Before Time 0, N (%)** |  |  |  |
| *Hospitalizations* |  |  |  |
| 0 | 34,682 (70.37%) | 34,074 (69.14%) | 0.03 |
| 1 | 8,388 (17.02%) | 8,557 (17.36%) | 0.01 |
| 2+ | 6,215 (12.61%) | 6,654 (13.50%) | 0.03 |
| *Emergency department visits* |  |  |  |
| 0 | 44,621 (90.54%) | 43,909 (89.09%) | 0.05 |
| 1 | 3,803 (7.72%) | 4,308 (8.74%) | 0.04 |
| 2+ | 861 (1.75%) | 1,068 (2.17%) | 0.03 |
| Skilled nursing facility stay | 31 (0.06%) | 36 (0.07%) | 0.00 |
| Influenza vaccination | 16,343 (33.16%) | 16,343 (33.16%) | 0.00 |
| Pneumococcal vaccination | 568 (1.15%) | 538 (1.09%) | 0.01 |
| Encounter for cancer screening | 13,424 (27.24%) | 12,875 (26.12%) | 0.03 |
| Eye examination | 2,976 (6.04%) | 2,837 (5.76%) | 0.01 |
| Colonoscopy | 2,006 (4.07%) | 1,981 (4.02%) | 0.00 |
| Bone mineral density test | 529 (1.07%) | 493 (1.00%) | 0.01 |
| Well-check/well-child preventive healthcare visit | 19,704 (39.98%) | 18,851 (38.25%) | 0.04 |
| Arthritis | 6,600 (13.39%) | 6,455 (13.10%) | 0.01 |
| Lipid abnormality | 8,711 (17.67%) | 8,650 (17.55%) | 0.00 |
| Ambulance use or life support services | 791 (1.60%) | 890 (1.81%) | 0.02 |
| Weakness | 941 (1.91%) | 924 (1.87%) | 0.00 |
| Pregnancy completion before Time 0 | 568 (2.65%) | 856 (3.99%) | 0.08 |
| **Characteristics Assessed Using all Available Data, N (%)** |  |  |  |
| Autoimmune disorders | 2,186 (4.44%) | 1,994 (4.05%) | 0.02 |
| Cancer | 2,822 (5.73%) | 2,622 (5.32%) | 0.02 |
| Chronic kidney disease or renal disease | 861 (1.75%) | 929 (1.88%) | 0.01 |
| Chronic liver disease | 1,943 (3.94%) | 2,068 (4.20%) | 0.01 |
| Chronic lung diseases (e.g., asthma, COPD, cystic fibrosis, pulmonary embolism) | 5,011 (10.17%) | 5,031 (10.21%) | 0.00 |
| Dementia or other neurological conditions | 3,168 (6.43%) | 3,115 (6.32%) | 0.00 |
| Diabetes mellitus, type 1 or 2 | 3,400 (6.90%) | 3,489 (7.08%) | 0.01 |
| Down syndrome | < 11 | < 11 | 0.00 |
| Heart conditions (e.g., heart failure, coronary artery disease, arrhythmias) | 6,147 (12.47%) | 6,519 (13.23%) | 0.02 |
| Hypertension | 10,468 (21.24%) | 10,757 (21.83%) | 0.01 |
| Immunocompromised state | 1,865 (3.78%) | 1,865 (3.78%) | 0.00 |
| Mental health conditions | 13,612 (27.62%) | 13,198 (26.78%) | 0.02 |
| Obese or severely obese | 9,764 (19.81%) | 9,928 (20.14%) | 0.01 |
| Sickle cell disease or thalassemia | 63 (0.13%) | 122 (0.25%) | 0.03 |
| Stroke or cerebrovascular disease | 714 (1.45%) | 822 (1.67%) | 0.02 |
| Tuberculosis | 23 (0.05%) | 24 (0.05%) | 0.00 |
| At least 1 COVID-19 laboratory performed | 17,668 (35.85%) | 16,423 (33.32%) | 0.05 |
| COVID-19 diagnoses occurring outside of a hospital or emergency department | 2,761 (5.60%) | 2,749 (5.58%) | 0.00 |
| Hospitalization or emergency department-diagnosed COVID-19 | 393 (0.80%) | 440 (0.89%) | 0.01 |

ASD = absolute standardized difference; COVID-19 = coronavirus disease 2019; Q1, Q3 = first and third quartiles; SD = standard deviation.
Note: Privacy rules require masking cell sizes of fewer than 11 individuals.

B. CVS Health
| Characteristic | Individuals vaccinated with JNJ-7836735<br>N = 149,813 | Matched unvaccinated individuals<br>N = 149,813 | ASD |
| --- | --- | --- | --- |
| **Characteristics assessed at Time 0** |  |  |  |
| *Age, years* |  |  |  |
| Median (Q1, Q3) | 44 (32, 54) | 44 (32, 54) | 0.00 |
| Mean (SD) | 42.98 (13.31) | 42.97 (13.32) | 0.00 |
| *Sex, N (%)* |  |  |  |
| Male | 82,697 (55.20%) | 82,697 (55.20%) | 0.00 |
| Female | 67,116 (44.80%) | 67,116 (44.80%) | 0.00 |
| *Region, N (%)* |  |  |  |
| Northeast | 29,728 (19.84%) | 29,728 (19.84%) | 0.00 |
| South | 26,237 (17.51%) | 26,237 (17.51%) | 0.00 |
| Midwest | 28,431 (18.98%) | 28,431 (18.98%) | 0.00 |
| West | 65,417 (43.67%) | 65,417 (43.67%) | 0.00 |
| Pregnant at Time 0, N (%) | 274 (0.18%) | 274 (0.18%) | 0.00 |
| **Characteristics in the 365 Days Before Time 0, N (%)** |  |  |  |
| *Hospitalizations* |  |  |  |
| 0 | 110,435 (73.72%) | 108,225 (72.24%) | 0.03 |
| 1 | 21,706 (14.49%) | 22,499 (15.02%) | 0.01 |
| 2+ | 17,672 (11.80%) | 19,089 (12.74%) | 0.03 |
| *Emergency department visits* |  |  |  |
| 0 | 136,145 (90.88%) | 134,052 (89.48%) | 0.05 |
| 1 | 11,123 (7.42%) | 12,310 (8.22%) | 0.03 |
| 2+ | 2,545 (1.70%) | 3,451 (2.30%) | 0.04 |
| Skilled nursing facility stay | 110 (0.07%) | 103 (0.07%) | 0.00 |
| Influenza vaccination | 44,501 (29.70%) | 44,501 (29.70%) | 0.00 |
| Pneumococcal vaccination | 1,675 (1.12%) | 1,661 (1.11%) | 0.00 |
| Encounter for cancer screening | 37,861 (25.27%) | 36,403 (24.30%) | 0.02 |
| Eye examination | 10,930 (7.30%) | 10,443 (6.97%) | 0.01 |
| Colonoscopy | 6,043 (4.03%) | 5,801 (3.87%) | 0.01 |
| Bone mineral density test | 1,839 (1.23%) | 1,778 (1.19%) | 0.00 |
| Well-check/well-child preventive healthcare visit | 54,304 (36.25%) | 51,984 (34.70%) | 0.03 |
| Arthritis | 21,142 (14.11%) | 20,700 (13.82%) | 0.01 |
| Lipid abnormality | 28,124 (18.77%) | 28,063 (18.73%) | 0.00 |
| Ambulance use or life support services | 2,439 (1.63%) | 2,882 (1.92%) | 0.02 |
| Weakness | 2,689 (1.79%) | 2,676 (1.79%) | 0.00 |
| Pregnancy completion before Time 0 | 1,703 (1.14%) | 2,198 (1.47%) | 0.03 |
| **Characteristics Assessed Using all Available Data, N (%)** |  |  |  |
| Autoimmune disorders | 6,805 (4.54%) | 6,812 (4.55%) | 0.00 |
| Cancer | 8,817 (5.89%) | 8,438 (5.63%) | 0.01 |
| Chronic kidney disease or renal disease | 3,300 (2.20%) | 3,559 (2.38%) | 0.01 |
| Chronic liver disease | 6,946 (4.64%) | 7,437 (4.96%) | 0.02 |
| Chronic lung diseases (e.g., asthma, COPD, cystic fibrosis, pulmonary embolism) | 16,433 (10.97%) | 16,778 (11.20%) | 0.01 |
| Dementia or other neurological conditions | 10,781 (7.20%) | 10,868 (7.25%) | 0.00 |
| Diabetes mellitus, type 1 or 2 | 10,737 (7.17%) | 11,694 (7.81%) | 0.02 |
| Down syndrome | 51 (0.03%) | 29 (0.02%) | 0.01 |
| Heart conditions (e.g., heart failure, coronary artery disease, arrhythmias) | 20,073 (13.40%) | 21,716 (14.50%) | 0.03 |
| Hypertension | 31,615 (21.10%) | 33,133 (22.12%) | 0.02 |
| Immunocompromised state | 5,561 (3.71%) | 5,561 (3.71%) | 0.00 |
| Mental health conditions | 42,143 (28.13%) | 41,064 (27.41%) | 0.02 |
| Obese or severely obese | 30,537 (20.38%) | 31,435 (20.98%) | 0.01 |
| Sickle cell disease or thalassemia | 359 (0.24%) | 415 (0.28%) | 0.01 |
| Stroke or cerebrovascular disease | 2,563 (1.71%) | 3,018 (2.01%) | 0.02 |
| Tuberculosis | 98 (0.07%) | 131 (0.09%) | 0.01 |
| At least 1 COVID-19 laboratory performed | 67,266 (44.90%) | 61,505 (41.05%) | 0.08 |
| COVID-19 diagnoses occurring outside of a hospital or emergency department | 7,207 (4.81%) | 7,095 (4.74%) | 0.00 |
| Hospitalization or emergency department-diagnosed COVID-19 | 1,076 (0.72%) | 1,271 (0.85%) | 0.01 |

ASD = absolute standardized difference; COVID-19 = coronavirus disease 2019; Q1, Q3 = first and third quartiles; SD = standard deviation.

**sTable 6.** Estimated Effectiveness of Receiving a Complete Primary Series of COVID-19 Vaccine in Adults Aged 18-64 Years, Compared With Being Unvaccinated, Overall.

| COVID-19 outcome | Vaccine exposure group | N | Events | Person-time (days) | HR (95% CI) | VE (95% CI) |
| --- | --- | --- | --- | --- | --- | --- |
| **Optum** |  |  |  |  |  |  |
| Medically diagnosed | BNT162b2 | 341,097 | 11,399 | 68,335,369 | 0.44 (0.42-0.45) | 56% (55%-58%) |
|  | Unvaccinated | 341,097 | 15,007 | 41,954,661 | — | — |
|  | mRNA-1273 | 201,604 | 5,691 | 42,145,941 | 0.34 (0.33-0.36) | 66% (64%-67%) |
|  | Unvaccinated | 201,604 | 9,122 | 25,093,307 | — | — |
|  | JNJ-7836735 | 49,285 | 2,215 | 10,249,729 | 0.56 (0.53-0.60) | 44% (40%-47%) |
|  | Unvaccinated | 49,285 | 2,327 | 6,337,889 | — | — |
| Hospital/ED-diagnosed | BNT162b2 | 341,097 | 1,066 | 69,030,682 | 0.18 (0.17-0.20) | 82% (80%-83%) |
|  | Unvaccinated | 341,097 | 3,470 | 42,860,899 | — | — |
|  | mRNA-1273 | 201,604 | 494 | 42,501,963 | 0.13 (0.12-0.14) | 87% (86%-88%) |
|  | Unvaccinated | 201,604 | 2,131 | 25,680,890 | — | — |
|  | JNJ-7836735 | 49,285 | 274 | 10,387,121 | 0.31 (0.26-0.36) | 69% (64%-74%) |
|  | Unvaccinated | 49,285 | 548 | 6,480,273 | — | — |
| **CVS Health** |  |  |  |  |  |  |
| Medically diagnosed | BNT162b2 | 1,151,775 | 40,116 | 268,538,241 | 0.51 (0.5-0.52) | 49% (48%-50%) |
|  | Unvaccinated | 1,151,775 | 47,292 | 167,925,461 | — | — |
|  | mRNA-1273 | 651,545 | 19,347 | 153,442,531 | 0.43 (0.42-0.44) | 57% (56%-58%) |
|  | Unvaccinated | 651,545 | 27,079 | 95,983,285 | — | — |
|  | JNJ-7836735 | 149,813 | 6,931 | 36,342,423 | 0.64 (0.62-0.66) | 36% (34%-38%) |
|  | Unvaccinated | 149,813 | 6,507 | 22,691,059 | — | — |
| Hospital/ED-diagnosed | BNT162b2 | 1,151,775 | 4,496 | 272,234,682 | 0.24 (0.23-0.25) | 76% (75%-77%) |
|  | Unvaccinated | 1,151,775 | 11,464 | 172,260,575 | — | — |
|  | mRNA-1273 | 651,545 | 2,042 | 155,254,572 | 0.18 (0.17-0.19) | 82% (81%-83%) |
|  | Unvaccinated | 651,545 | 6,884 | 98,442,974 | — | — |
|  | JNJ-7836735 | 149,813 | 957 | 37,004,803 | 0.35 (0.32-0.38) | 65% (62%-68%) |
|  | Unvaccinated | 149,813 | 1,702 | 23,256,138 | — | — |

CI = confidence interval; COVID-19 = coronavirus disease 2019; ED = emergency department; HR = hazard ratio; VE = vaccine effectiveness.
Note: — indicates the reference group.

**sTable 7.** Estimated Effectiveness of Receiving a Complete Primary Series of COVID-19 Vaccine in Adults Aged 18-64 Years, Compared With Being Unvaccinated, by Previous COVID-19 Diagnosis Status.

| Subgroup | COVID-19 outcome | Vaccine exposure group | N | Events | Person-time (days) | HR (95% CI) | VE (95% CI) |
| --- | --- | --- | --- | --- | --- | --- | --- |
| **Optum** |  |  |  |  |  |  |  |
| With previous COVID-19 diagnosis<sup>a</sup> | Medically diagnosed | BNT162b2 | 18,962 | 759 | 3,389,905 | 0.66 (0.59-0.74) | 34% (26%-41%) |
|  |  | Unvaccinated | 18,962 | 779 | 2,334,640 | — | — |
|  |  | mRNA-1273 | 9,914 | 341 | 1,857,921 | 0.55 (0.47-0.64) | 45% (36%-53%) |
|  |  | Unvaccinated | 9,914 | 409 | 1,248,699 | — | — |
|  | Hospital/ED-diagnosed | BNT162b2 | 18,962 | 39 | 3,454,998 | 0.28 (0.18-0.43) | 72% (57%-82%) |
|  |  | Unvaccinated | 18,962 | 92 | 2,381,274 | — | — |
|  |  | mRNA-1273 | 9,914 | 16 | 1,886,452 | 0.25 (0.14-0.45) | 75% (55%-86%) |
|  |  | Unvaccinated | 9,914 | 42 | 1,280,414 | — | — |
| Without a previous COVID-19 diagnosis | Medically diagnosed | BNT162b2 | 322,135 | 10,640 | 64,945,464 | 0.42 (0.41-0.44) | 58% (56%-59%) |
|  |  | Unvaccinated | 322,135 | 14,228 | 39,620,021 | — | — |
|  |  | mRNA-1273 | 191,690 | 5,350 | 40,288,020 | 0.33 (0.32-0.35) | 67% (65%-68%) |
|  |  | Unvaccinated | 191,690 | 8,713 | 23,844,608 | — | — |
|  | Hospital/ED-diagnosed | BNT162b2 | 322,135 | 1,027 | 65,575,684 | 0.18 (0.16-0.19) | 82% (81%-84%) |
|  |  | Unvaccinated | 322,135 | 3,378 | 40,479,625 | — | — |
|  |  | mRNA-1273 | 191,690 | 478 | 40,615,511 | 0.13 (0.12-0.14) | 87% (86%-88%) |
|  |  | Unvaccinated | 191,690 | 2,089 | 24,400,476 | — | — |
| **CVS Health** |  |  |  |  |  |  |  |
| With previous COVID-19 diagnosis<sup>a</sup> | Medically diagnosed | BNT162b2 | 56,266 | 2,768 | 12,001,685 | 0.71 (0.67-0.75) | 29% (25%-33%) |
|  |  | Unvaccinated | 56,266 | 2,745 | 8,365,317 | — | — |
|  |  | mRNA-1273 | 29,029 | 1,350 | 6,297,879 | 0.66 (0.61-0.72) | 34% (28%-39%) |
|  |  | Unvaccinated | 29,029 | 1,436 | 4,371,147 | — | — |
|  |  | JNJ-7836735 | 7,660 | 406 | 1,743,282 | 0.74 (0.64-0.85) | 26% (15%-36%) |
|  |  | Unvaccinated | 7,660 | 371 | 1,164,613 | — | — |
|  | Hospital/ED-diagnosed | BNT162b2 | 56,266 | 186 | 12,324,343 | 0.44 (0.35-0.53) | 56% (47%-65%) |
|  |  | Unvaccinated | 56,266 | 306 | 8,644,775 | — | — |
|  |  | mRNA-1273 | 29,029 | 87 | 6,472,352 | 0.34 (0.26-0.45) | 66% (55%-74%) |
|  |  | Unvaccinated | 29,029 | 174 | 4,519,359 | — | — |
|  |  | JNJ-7836735 | 7,660 | 28 | 1,790,309 | 0.59 (0.36-0.99) | 41% (1%-64%) |
|  |  | Unvaccinated | 7,660 | 34 | 1,198,980 | — | — |
| Without a previous COVID-19 diagnosis | Medically diagnosed | BNT162b2 | 1,095,509 | 37,348 | 256,536,556 | 0.50 (0.49-0.50) | 50% (50%-51%) |
|  |  | Unvaccinated | 1,095,509 | 44,547 | 159,560,144 | — | — |
|  |  | mRNA-1273 | 622,516 | 17,997 | 147,144,652 | 0.42 (0.41-0.42) | 58% (58%-59%) |
|  |  | Unvaccinated | 622,516 | 25,643 | 91,612,138 | — | — |
|  |  | JNJ-7836735 | 142,153 | 6,525 | 34,599,141 | 0.63 (0.61-0.65) | 37% (35%-39%) |
|  |  | Unvaccinated | 142,153 | 6,136 | 21,526,446 | — | — |
|  | Hospital/ED-diagnosed | BNT162b2 | 1,095,509 | 4,310 | 259,910,339 | 0.24 (0.23-0.25) | 76% (75%-77%) |
|  |  | Unvaccinated | 1,095,509 | 11,158 | 163,615,800 | — | — |
|  |  | mRNA-1273 | 622,516 | 1,955 | 148,782,220 | 0.17 (0.17-0.18) | 83% (82%-83%) |
|  |  | Unvaccinated | 622,516 | 6,710 | 93,923,615 | — | — |
|  |  | JNJ-7836735 | 142,153 | 929 | 35,214,494 | 0.34 (0.32-0.37) | 66% (63%-68%) |
|  |  | Unvaccinated | 142,153 | 1,668 | 22,057,158 | — | — |

CI = confidence interval; COVID-19 = coronavirus disease 2019; ED = emergency department; HR = hazard ratio; VE = vaccine effectiveness.
Note: — indicates the reference group.
<sup>a</sup> Having a diagnosis of COVID-19 in any medically attended setting at any point before Time 0.

**sTable 8.** Estimated Effectiveness of Receiving a Complete Primary Series of COVID-19 Vaccine Compared with Being Unvaccinated, Corrected for Potentially Missing Vaccine Records.

| COVID-19 outcome | Vaccine | Hypothesized sensitivity of vaccination exposure measurement | Corrected VE (95% CI) |
| --- | --- | --- | --- |
| Optum |  |  |  |
| Medically diagnosed | BNT162b2 | 100% (uncorrected) | 56% (55%-58%) |
|  |  | 83% | 61% (60%-63%) |
|  |  | 71% | 67% (66%-68%) |
|  | mRNA-1273 | 100% (uncorrected) | 66% (64%-67%) |
|  |  | 83% | 70% (68%-71%) |
|  |  | 71% | 75% (74%-76%) |
|  | JNJ-7836735 | 100% (uncorrected) | 44% (40%-47%) |
|  |  | 83% | 49% (45%-52%) |
|  |  | 71% | 56% (53%-58%) |
| Hospital/ED-diagnosed | BNT162b2 | 100% (uncorrected) | 82% (80%-83%) |
|  |  | 83% | 85% (83%-85%) |
|  |  | 71% | 88% (86%-88%) |
|  | mRNA-1273 | 100% (uncorrected) | 87% (86%-88%) |
|  |  | 83% | 89% (88%-90%) |
|  |  | 71% | 91% (91%-92%) |
|  | JNJ-7836735 | 100% (uncorrected) | 69% (64%-74%) |
|  |  | 83% | 73% (69%-77%) |
|  |  | 71% | 78% (74%-81%) |
| CVS Health |  |  |  |
| Medically diagnosed | BNT162b2 | 100% (uncorrected) | 49% (48%-50%) |
|  |  | 89% | 52% (52%-53%) |
|  |  | 69% | 63% (62%-64%) |
|  | mRNA-1273 | 100% (uncorrected) | 57% (56%-58%) |
|  |  | 89% | 60% (59%-61%) |
|  |  | 69% | 69% (69%-70%) |
|  | JNJ-7836735 | 100% (uncorrected) | 36% (34%-38%) |
|  |  | 89% | 38% (36%-40%) |
|  |  | 69% | 46% (44%-48%) |
| Hospital/ED-diagnosed | BNT162b2 | 100% (uncorrected) | 76% (75%-77%) |
|  |  | 89% | 78% (77%-79%) |
|  |  | 69% | 84% (83%-85%) |
|  | mRNA-1273 | 100% (uncorrected) | 82% (81%-83%) |
|  |  | 89% | 84% (83%-85%) |
|  |  | 69% | 89% (88%-89%) |
|  | JNJ-7836735 | 100% (uncorrected) | 65% (62%-68%) |
|  |  | 89% | 67% (64%-70%) |
|  |  | 69% | 74% (72%-76%) |

CI = confidence interval; COVID-19 = coronavirus disease 2019; VE = vaccine effectiveness.

**sTable 9.**
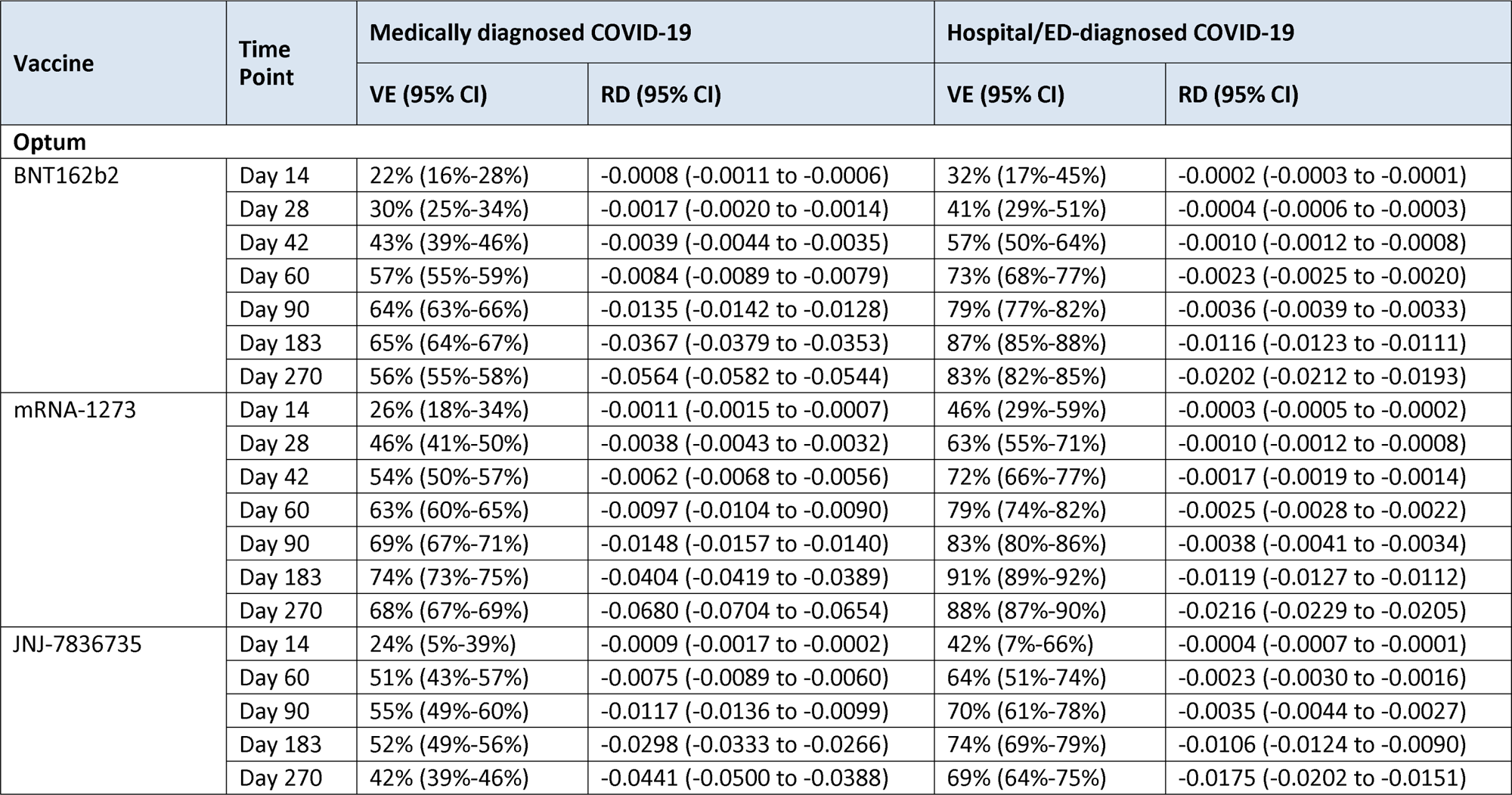
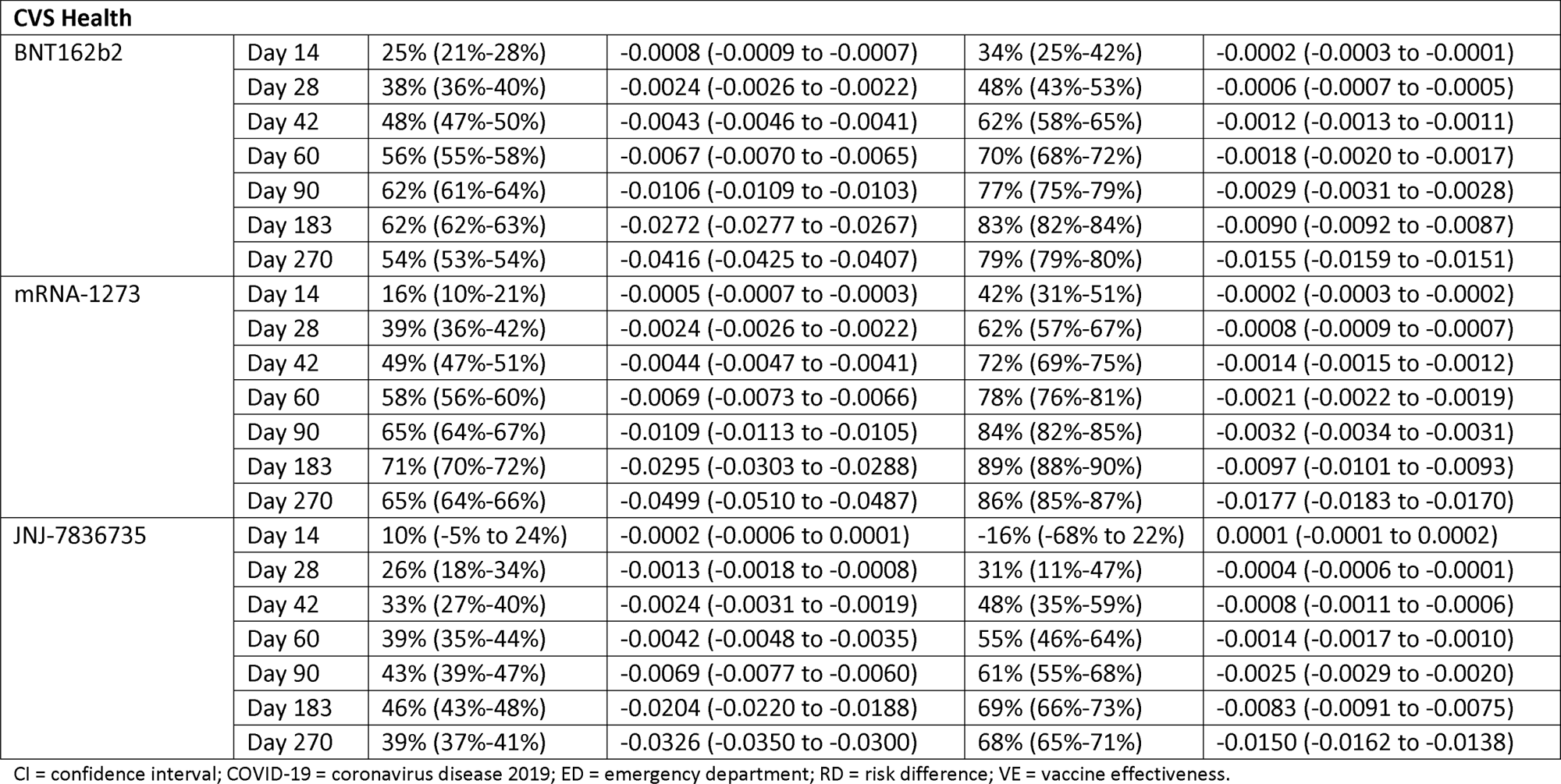
Estimated Effectiveness of Receiving a Complete Primary Series of COVID-19 Vaccine Compared With Being Unvaccinated, Over Time.

| Vaccine | Time Point | Medically diagnosed COVID-19 |  | Hospital/ED-diagnosed COVID-19 |  |
| --- | --- | --- | --- | --- | --- |
|  |  | VE (95% CI) | RD (95% CI) | VE (95% CI) | RD (95% CI) |
| Optum |  |  |  |  |  |
| BNT162b2 | Day 14 | 22% (16%-28%) | -0.0008 (-0.0011 to -0.0006) | 32% (17%-45%) | -0.0002 (-0.0003 to -0.0001) |
|  | Day 28 | 30% (25%-34%) | -0.0017 (-0.0020 to -0.0014) | 41% (29%-51%) | -0.0004 (-0.0006 to -0.0003) |
|  | Day 42 | 43% (39%-46%) | -0.0039 (-0.0044 to -0.0035) | 57% (50%-64%) | -0.0010 (-0.0012 to -0.0008) |
|  | Day 60 | 57% (55%-59%) | -0.0084 (-0.0089 to -0.0079) | 73% (68%-77%) | -0.0023 (-0.0025 to -0.0020) |
|  | Day 90 | 64% (63%-66%) | -0.0135 (-0.0142 to -0.0128) | 79% (77%-82%) | -0.0036 (-0.0039 to -0.0033) |
|  | Day 183 | 65% (64%-67%) | -0.0367 (-0.0379 to -0.0353) | 87% (85%-88%) | -0.0116 (-0.0123 to -0.0111) |
|  | Day 270 | 56% (55%-58%) | -0.0564 (-0.0582 to -0.0544) | 83% (82%-85%) | -0.0202 (-0.0212 to -0.0193) |
| mRNA-1273 | Day 14 | 26% (18%-34%) | -0.0011 (-0.0015 to -0.0007) | 46% (29%-59%) | -0.0003 (-0.0005 to -0.0002) |
|  | Day 28 | 46% (41%-50%) | -0.0038 (-0.0043 to -0.0032) | 63% (55%-71%) | -0.0010 (-0.0012 to -0.0008) |
|  | Day 42 | 54% (50%-57%) | -0.0062 (-0.0068 to -0.0056) | 72% (66%-77%) | -0.0017 (-0.0019 to -0.0014) |
|  | Day 60 | 63% (60%-65%) | -0.0097 (-0.0104 to -0.0090) | 79% (74%-82%) | -0.0025 (-0.0028 to -0.0022) |
|  | Day 90 | 69% (67%-71%) | -0.0148 (-0.0157 to -0.0140) | 83% (80%-86%) | -0.0038 (-0.0041 to -0.0034) |
|  | Day 183 | 74% (73%-75%) | -0.0404 (-0.0419 to -0.0389) | 91% (89%-92%) | -0.0119 (-0.0127 to -0.0112) |
|  | Day 270 | 68% (67%-69%) | -0.0680 (-0.0704 to -0.0654) | 88% (87%-90%) | -0.0216 (-0.0229 to -0.0205) |
| JNJ-7836735 | Day 14 | 24% (5%-39%) | -0.0009 (-0.0017 to -0.0002) | 42% (7%-66%) | -0.0004 (-0.0007 to -0.0001) |
|  | Day 60 | 51% (43%-57%) | -0.0075 (-0.0089 to -0.0060) | 64% (51%-74%) | -0.0023 (-0.0030 to -0.0016) |
|  | Day 90 | 55% (49%-60%) | -0.0117 (-0.0136 to -0.0099) | 70% (61%-78%) | -0.0035 (-0.0044 to -0.0027) |
|  | Day 183 | 52% (49%-56%) | -0.0298 (-0.0333 to -0.0266) | 74% (69%-79%) | -0.0106 (-0.0124 to -0.0090) |
|  | Day 270 | 42% (39%-46%) | -0.0441 (-0.0500 to -0.0388) | 69% (64%-75%) | -0.0175 (-0.0202 to -0.0151) |
|  |  | VE (95% CI) | RD (95% CI) | VE (95% CI) | RD (95% CI) |
| CVS Health |  |  |  |  |  |
| BNT162b2 | Day 14 | 25% (21%-28%) | -0.0008 (-0.0009 to -0.0007) | 34% (25%-42%) | -0.0002 (-0.0003 to -0.0001) |
|  | Day 28 | 38% (36%-40%) | -0.0024 (-0.0026 to -0.0022) | 48% (43%-53%) | -0.0006 (-0.0007 to -0.0005) |
|  | Day 42 | 48% (47%-50%) | -0.0043 (-0.0046 to -0.0041) | 62% (58%-65%) | -0.0012 (-0.0013 to -0.0011) |
|  | Day 60 | 56% (55%-58%) | -0.0067 (-0.0070 to -0.0065) | 70% (68%-72%) | -0.0018 (-0.0020 to -0.0017) |
|  | Day 90 | 62% (61%-64%) | -0.0106 (-0.0109 to -0.0103) | 77% (75%-79%) | -0.0029 (-0.0031 to -0.0028) |
|  | Day 183 | 62% (62%-63%) | -0.0272 (-0.0277 to -0.0267) | 83% (82%-84%) | -0.0090 (-0.0092 to -0.0087) |
|  | Day 270 | 54% (53%-54%) | -0.0416 (-0.0425 to -0.0407) | 79% (79%-80%) | -0.0155 (-0.0159 to -0.0151) |
| mRNA-1273 | Day 14 | 16% (10%-21%) | -0.0005 (-0.0007 to -0.0003) | 42% (31%-51%) | -0.0002 (-0.0003 to -0.0002) |
|  | Day 28 | 39% (36%-42%) | -0.0024 (-0.0026 to -0.0022) | 62% (57%-67%) | -0.0008 (-0.0009 to -0.0007) |
|  | Day 42 | 49% (47%-51%) | -0.0044 (-0.0047 to -0.0041) | 72% (69%-75%) | -0.0014 (-0.0015 to -0.0012) |
|  | Day 60 | 58% (56%-60%) | -0.0069 (-0.0073 to -0.0066) | 78% (76%-81%) | -0.0021 (-0.0022 to -0.0019) |
|  | Day 90 | 65% (64%-67%) | -0.0109 (-0.0113 to -0.0105) | 84% (82%-85%) | -0.0032 (-0.0034 to -0.0031) |
|  | Day 183 | 71% (70%-72%) | -0.0295 (-0.0303 to -0.0288) | 89% (88%-90%) | -0.0097 (-0.0101 to -0.0093) |
|  | Day 270 | 65% (64%-66%) | -0.0499 (-0.0510 to -0.0487) | 86% (85%-87%) | -0.0177 (-0.0183 to -0.0170) |
| JNJ-7836735 | Day 14 | 10% (-5% to 24%) | -0.0002 (-0.0006 to 0.0001) | -16% (-68% to 22%) | 0.0001 (-0.0001 to 0.0002) |
|  | Day 28 | 26% (18%-34%) | -0.0013 (-0.0018 to -0.0008) | 31% (11%-47%) | -0.0004 (-0.0006 to -0.0001) |
|  | Day 42 | 33% (27%-40%) | -0.0024 (-0.0031 to -0.0019) | 48% (35%-59%) | -0.0008 (-0.0011 to -0.0006) |
|  | Day 60 | 39% (35%-44%) | -0.0042 (-0.0048 to -0.0035) | 55% (46%-64%) | -0.0014 (-0.0017 to -0.0010) |
|  | Day 90 | 43% (39%-47%) | -0.0069 (-0.0077 to -0.0060) | 61% (55%-68%) | -0.0025 (-0.0029 to -0.0020) |
|  | Day 183 | 46% (43%-48%) | -0.0204 (-0.0220 to -0.0188) | 69% (66%-73%) | -0.0083 (-0.0091 to -0.0075) |
|  | Day 270 | 39% (37%-41%) | -0.0326 (-0.0350 to -0.0300) | 68% (65%-71%) | -0.0150 (-0.0162 to -0.0138) |

CI = confidence interval; COVID-19 = coronavirus disease 2019; ED = emergency department; RD = risk difference; VE = vaccine effectiveness.

**sFigure 1.**
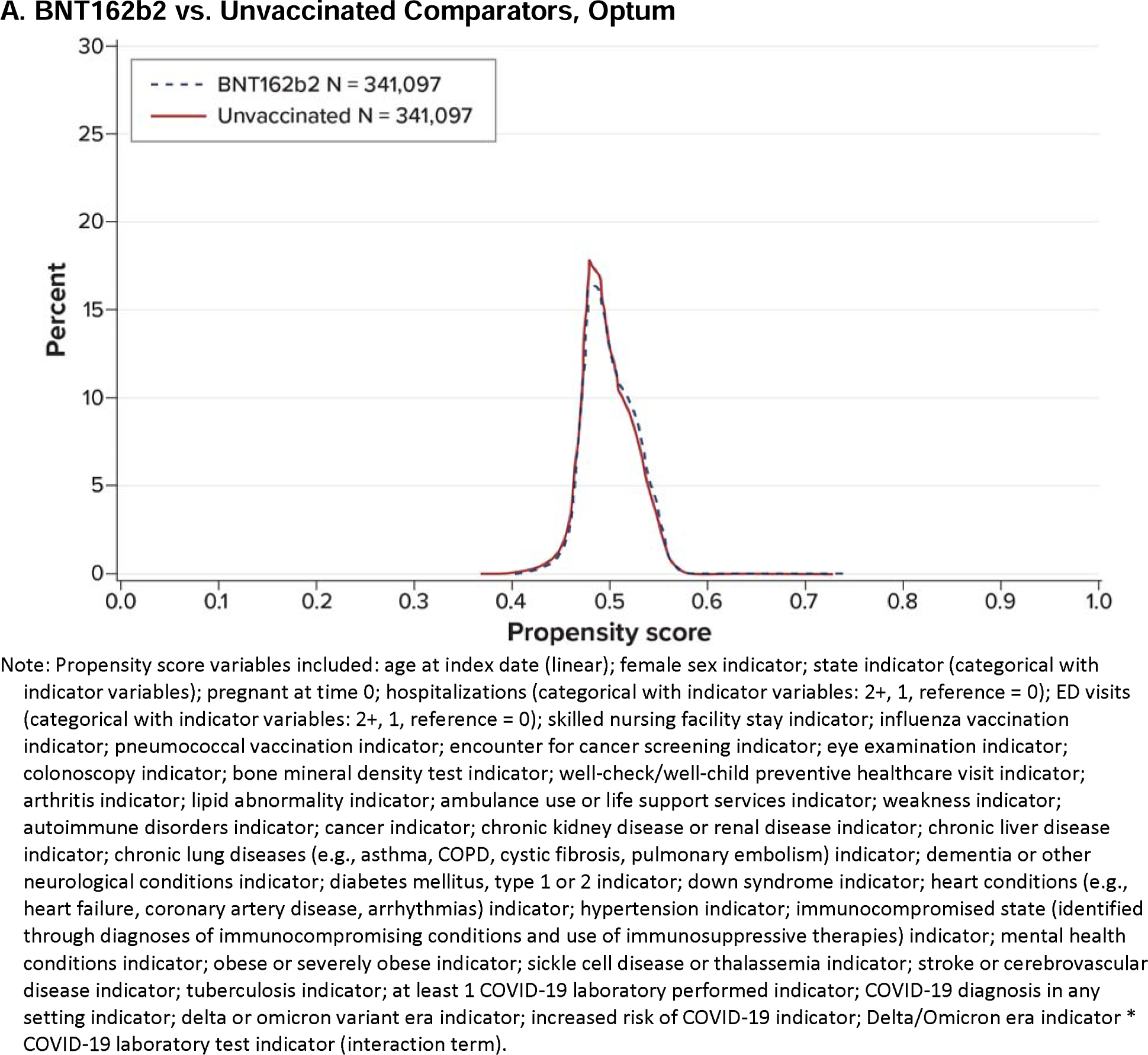
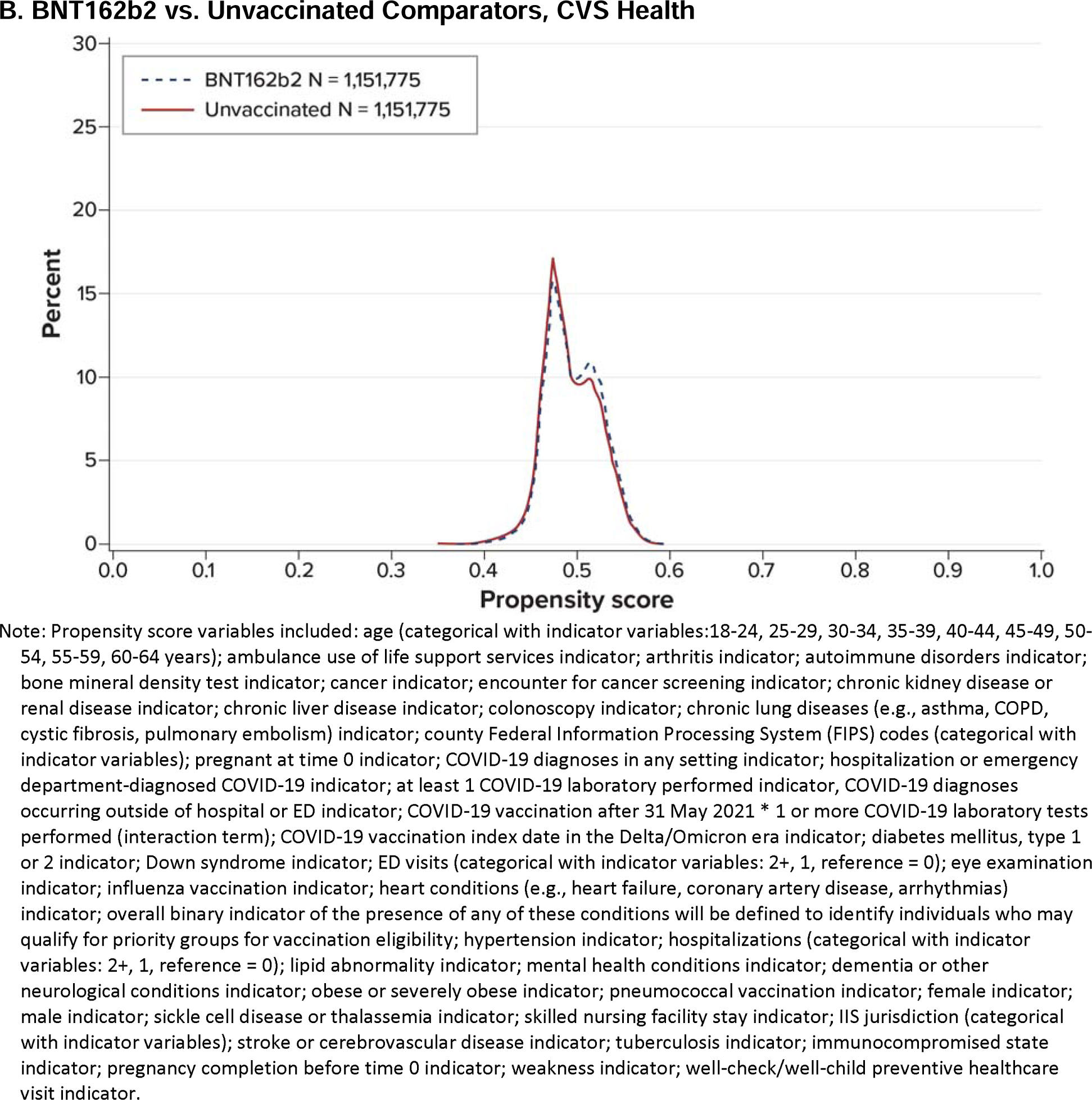
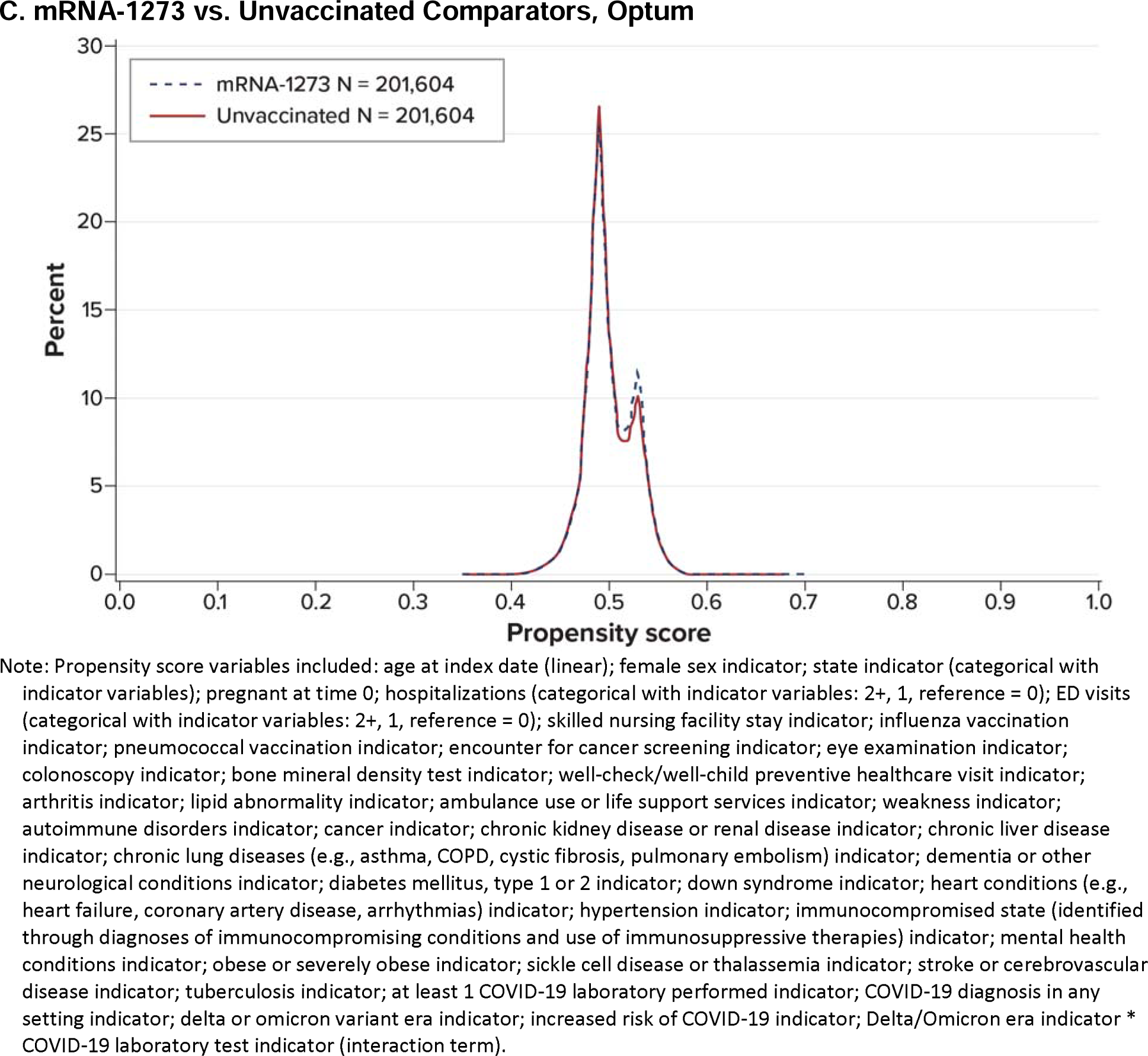
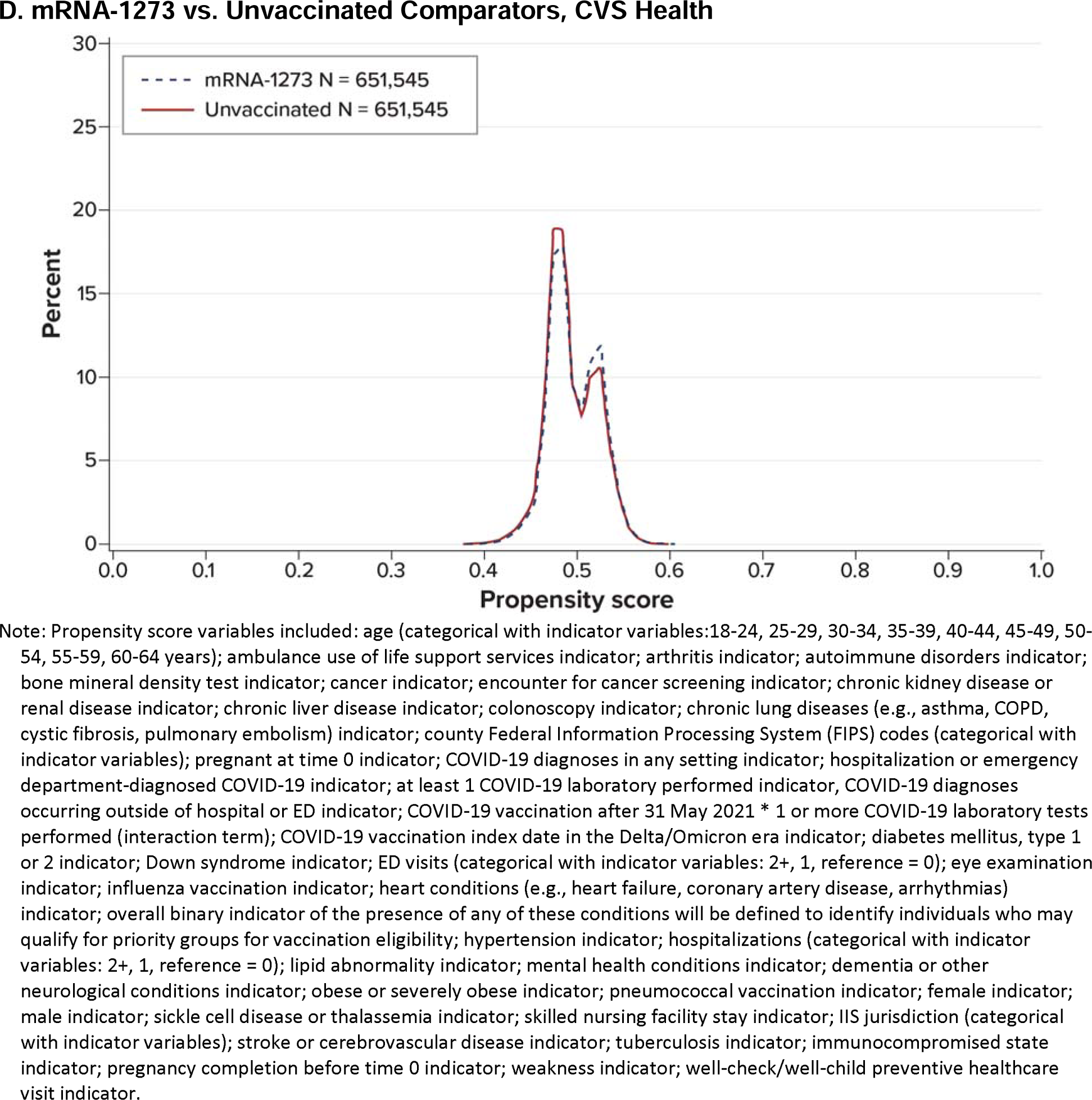
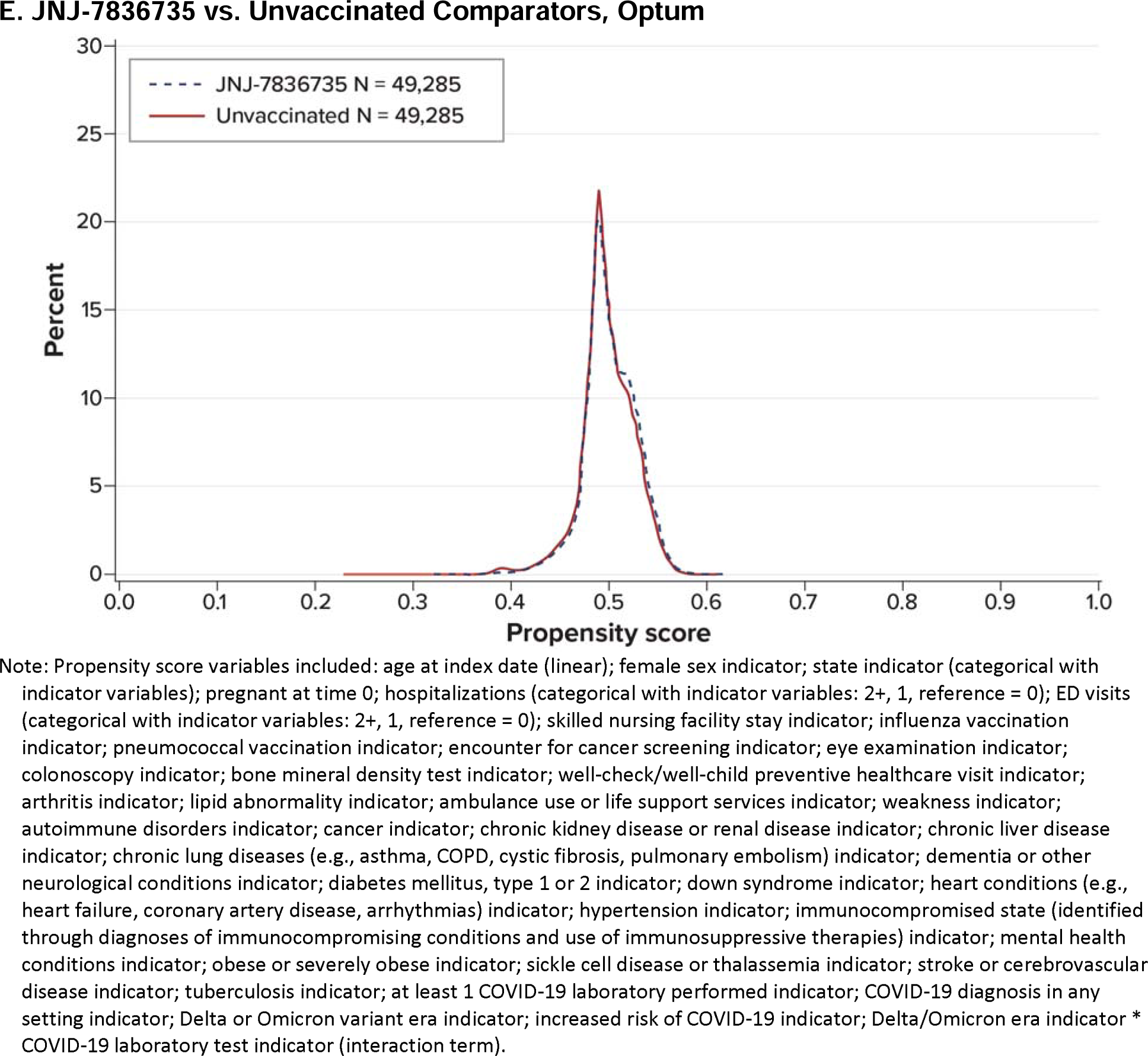
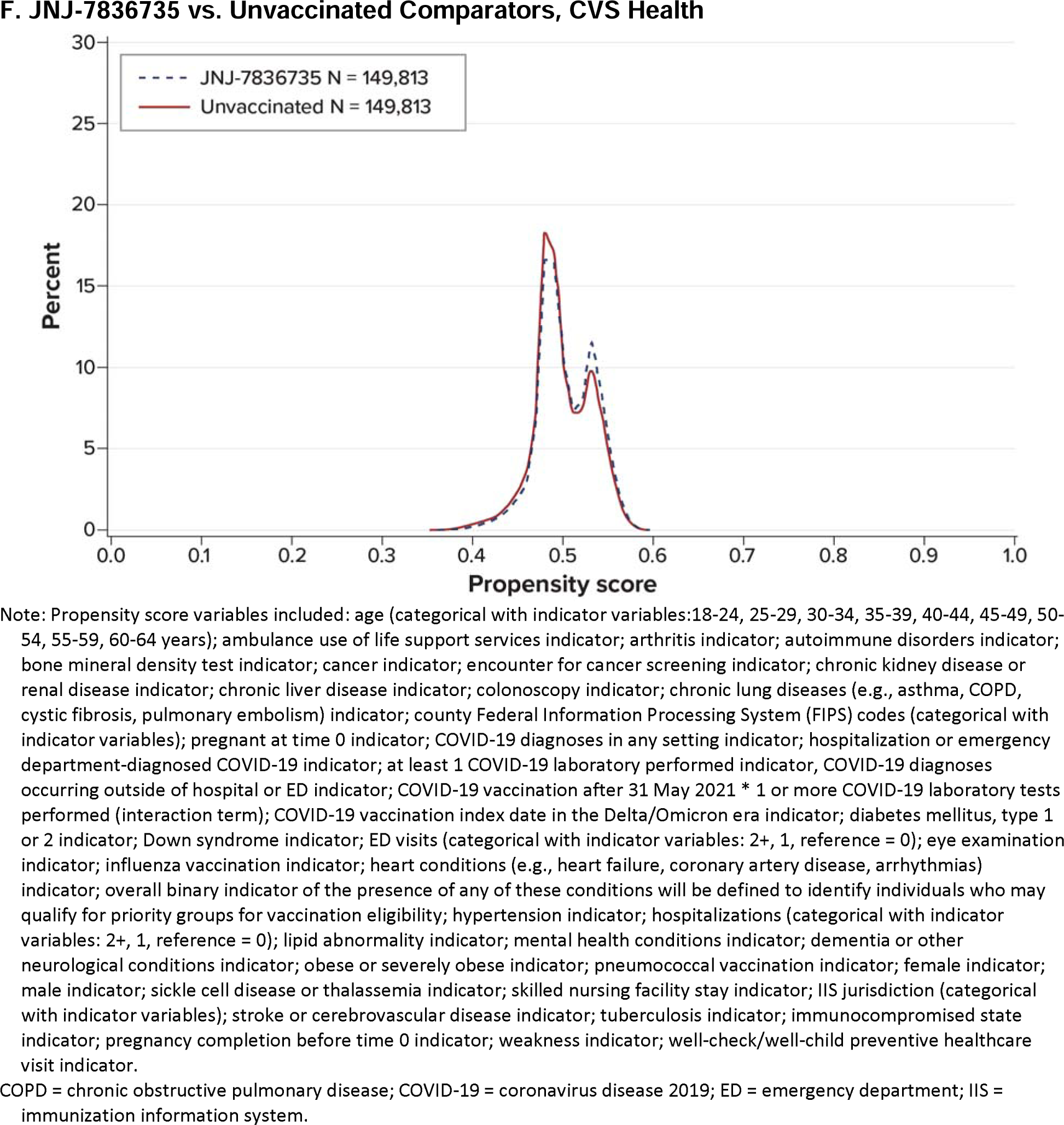
Propensity Score Distributions of Adults Aged 18-64 Years Receiving a Complete Primary Series of COVID-19 Vaccine and Matched Unvaccinated Comparators.

**sFigure 2.**
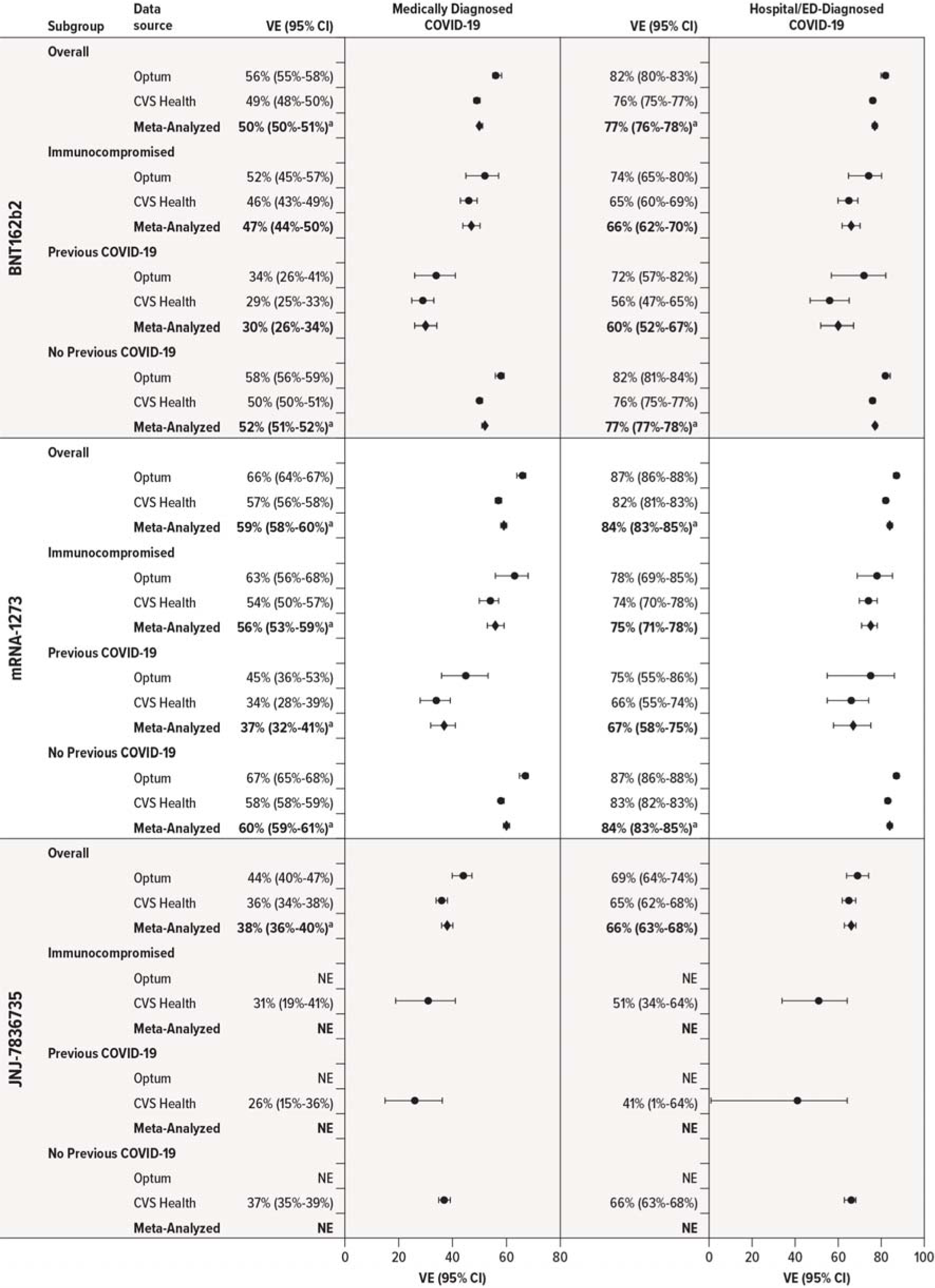
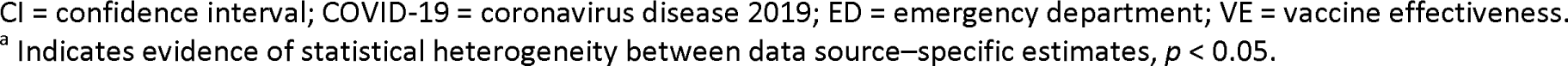
Estimated Effectiveness of Receiving a Complete Primary Series of COVID-19 Vaccine in Adults Aged 18-64 Years, Compared With Being Unvaccinated, Overall and Within Subgroups.

**sFigure 3.**
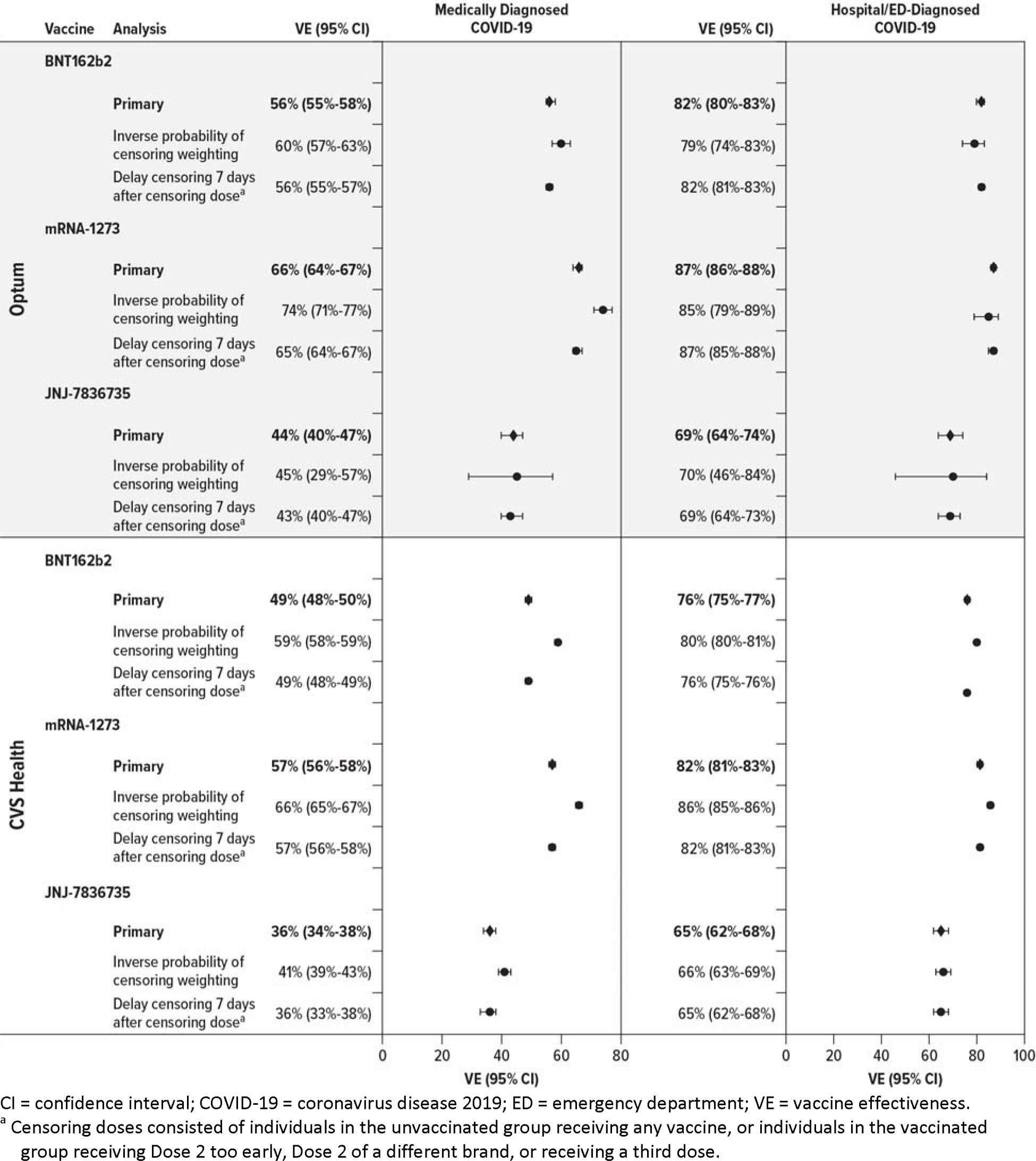
Estimated Effectiveness of Receiving a Complete Primary Series of COVID-19 Vaccine in Adults Aged 18-64 Years, Compared With Being Unvaccinated, Primary and Sensitivity Analyses.

**sFigure 4.**
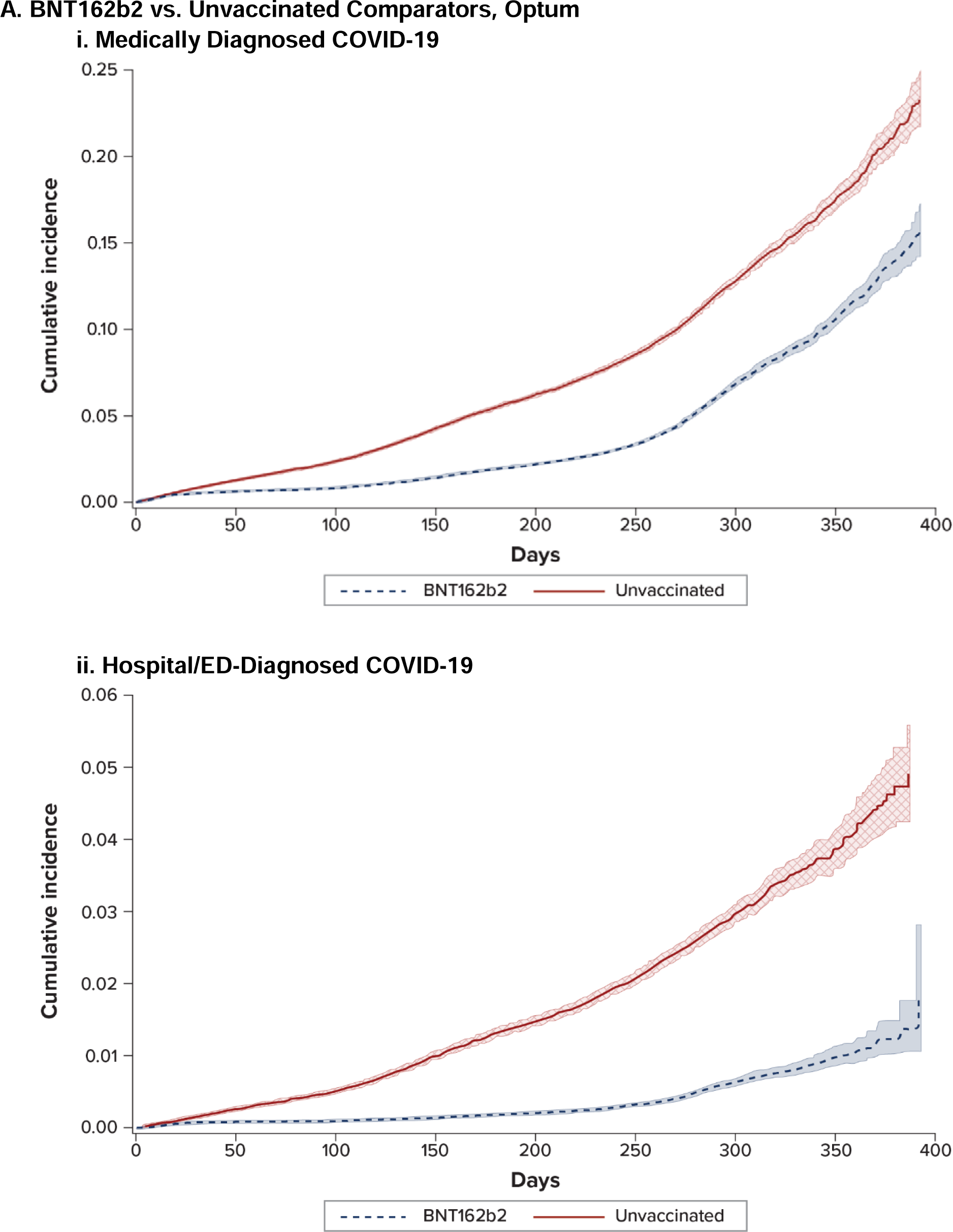
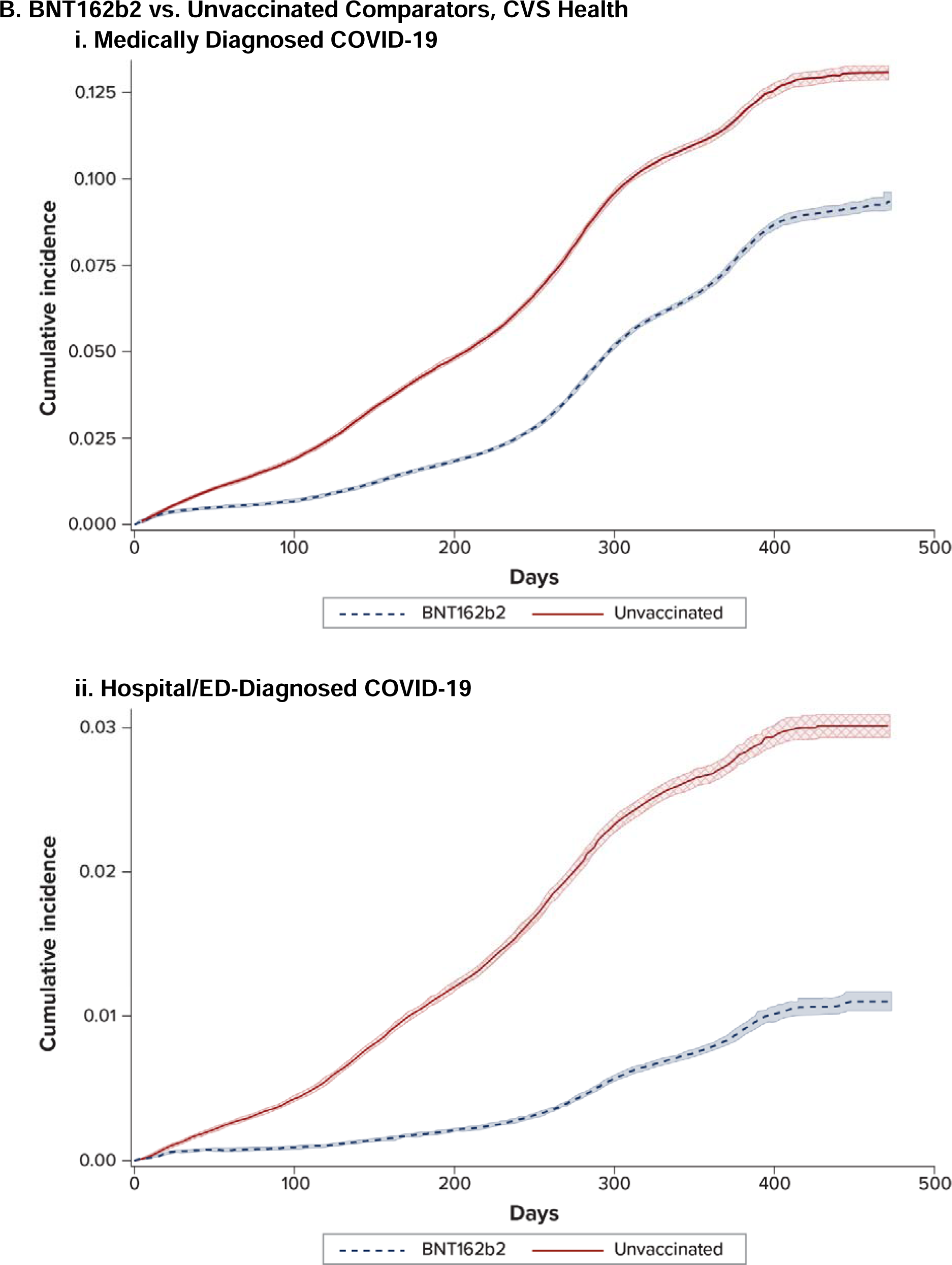
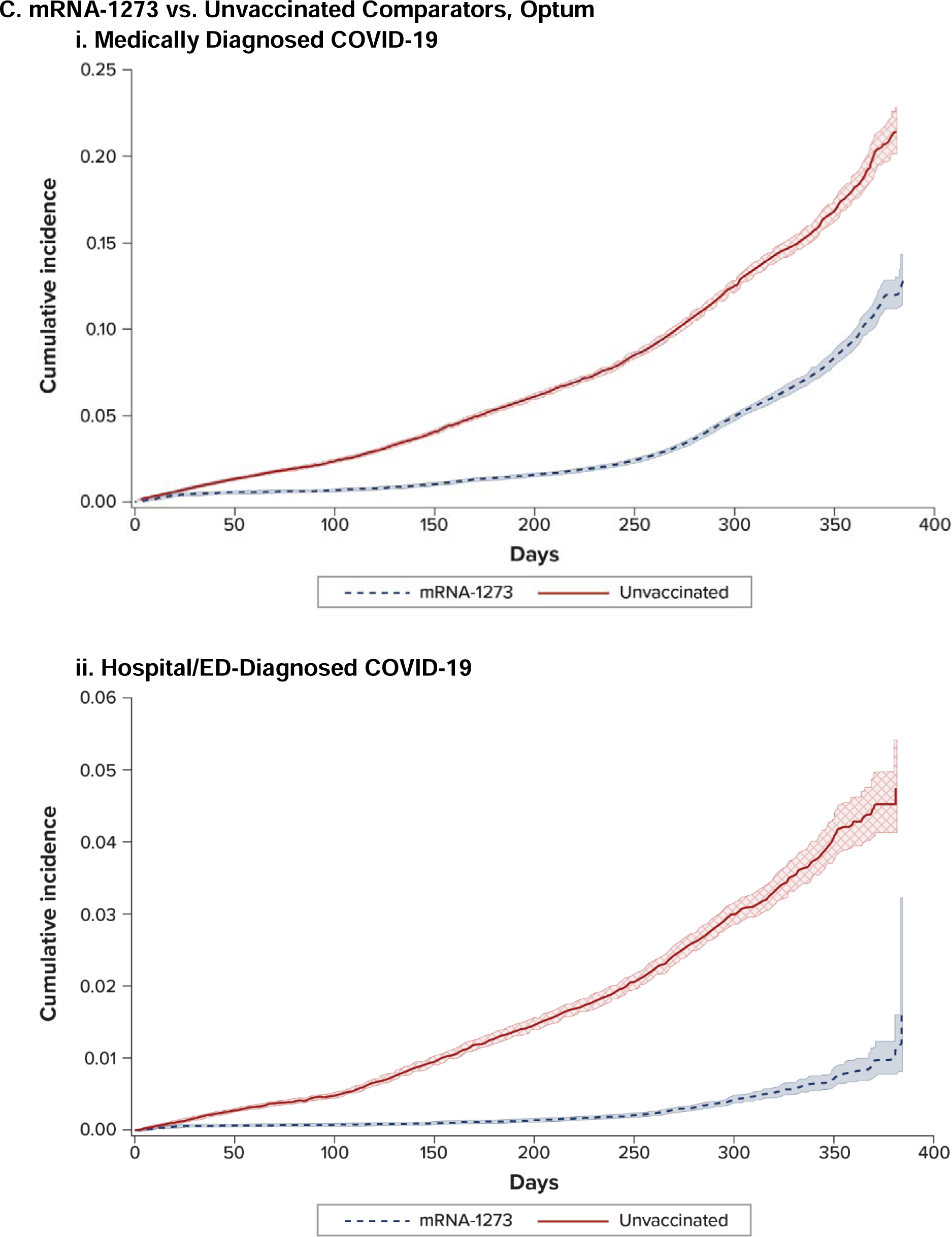
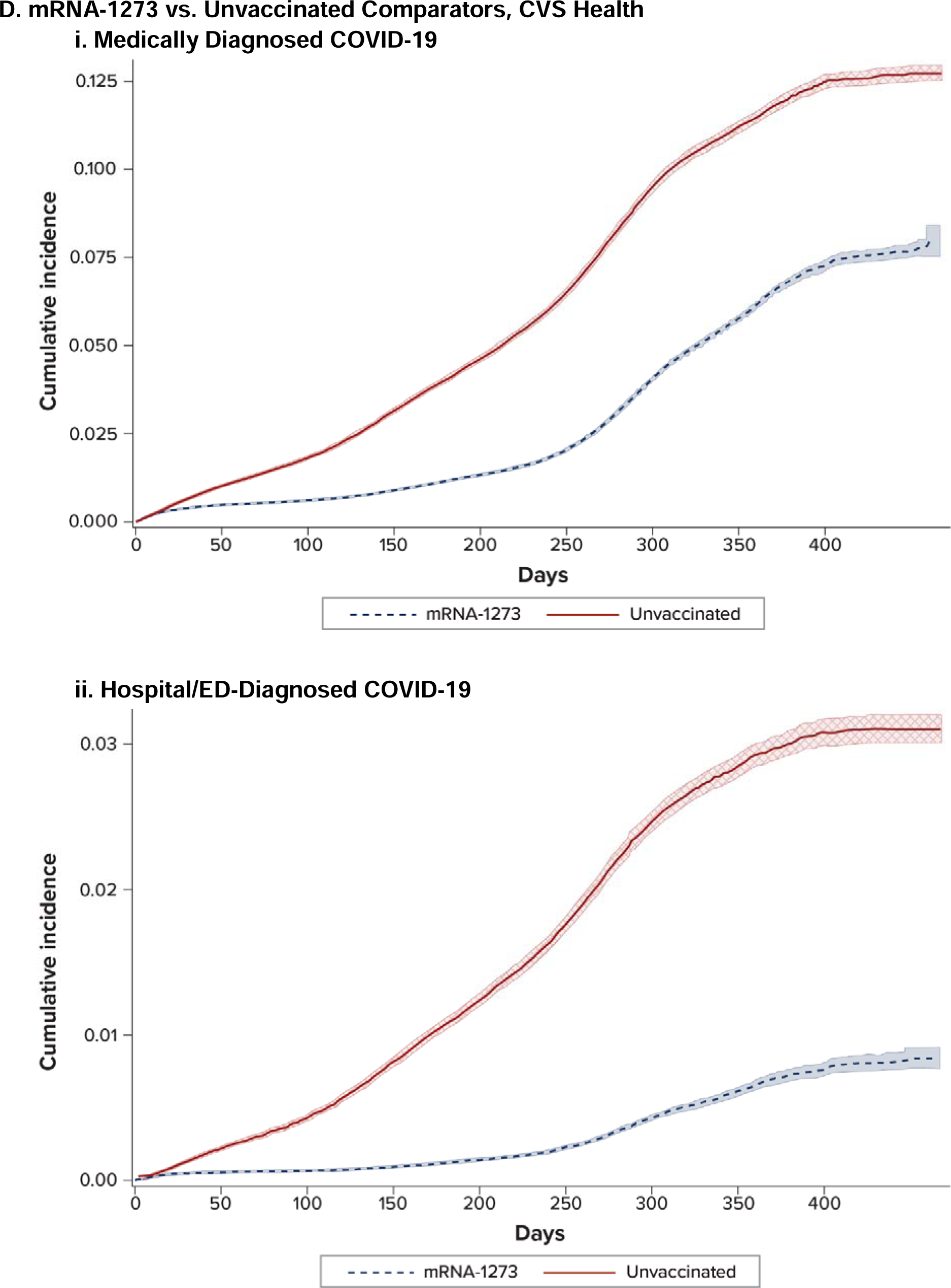
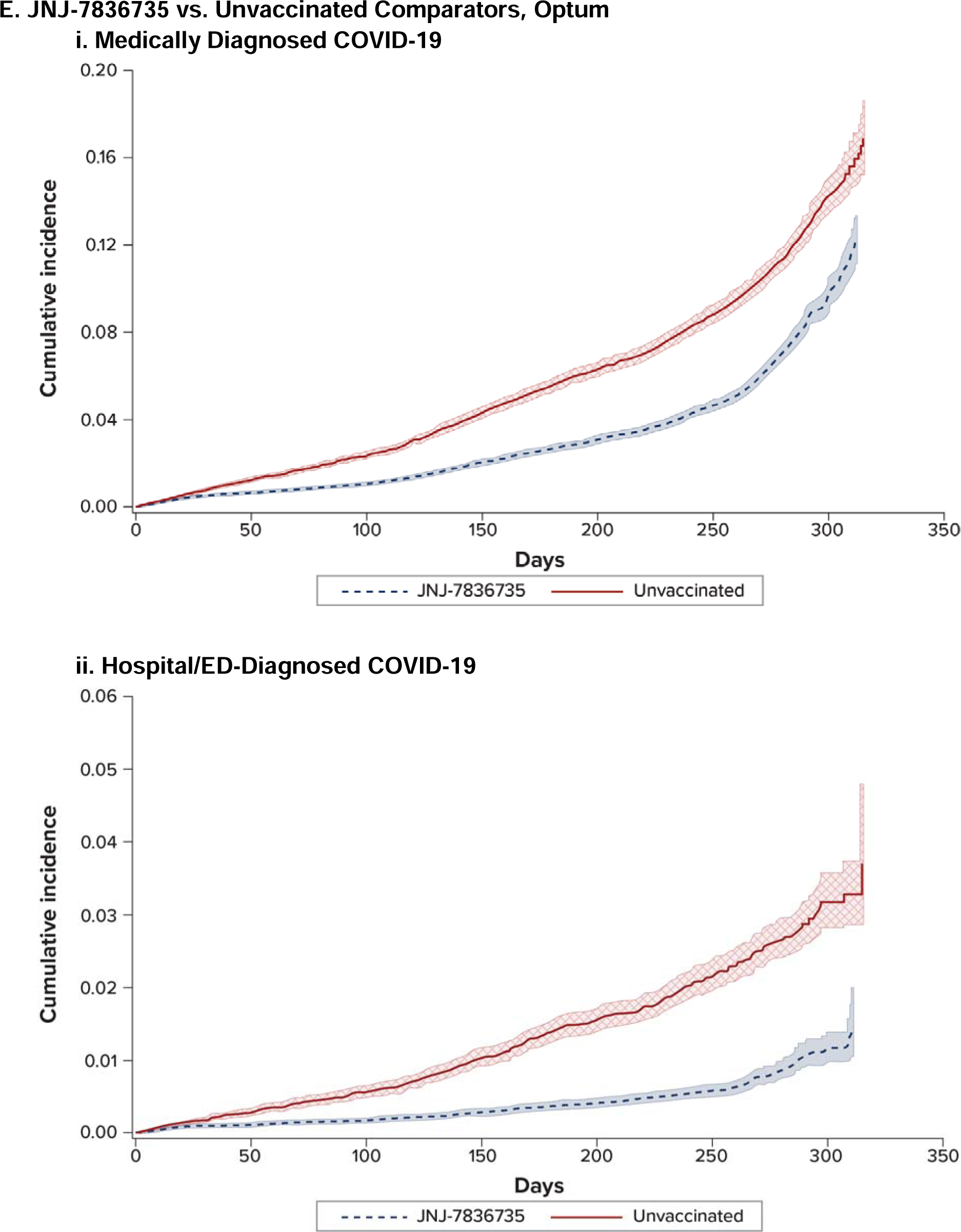
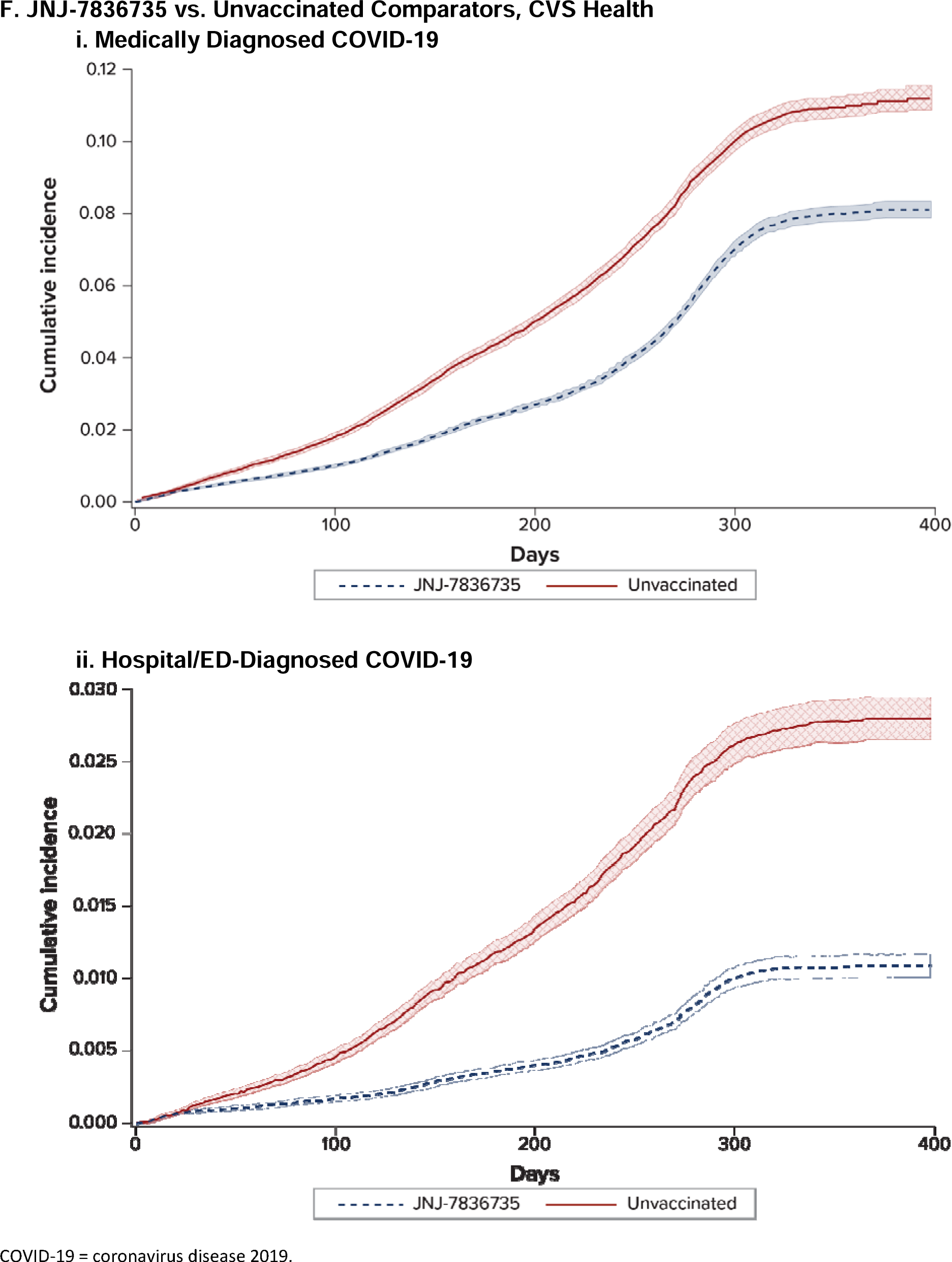
Weighted Cumulative Incidence of COVID-19 Outcomes in Adults Aged 18-64 Years Receiving a Complete Primary Series of COVID-19 Vaccines and Matched Unvaccinated Comparators.

**sFigure 5.**
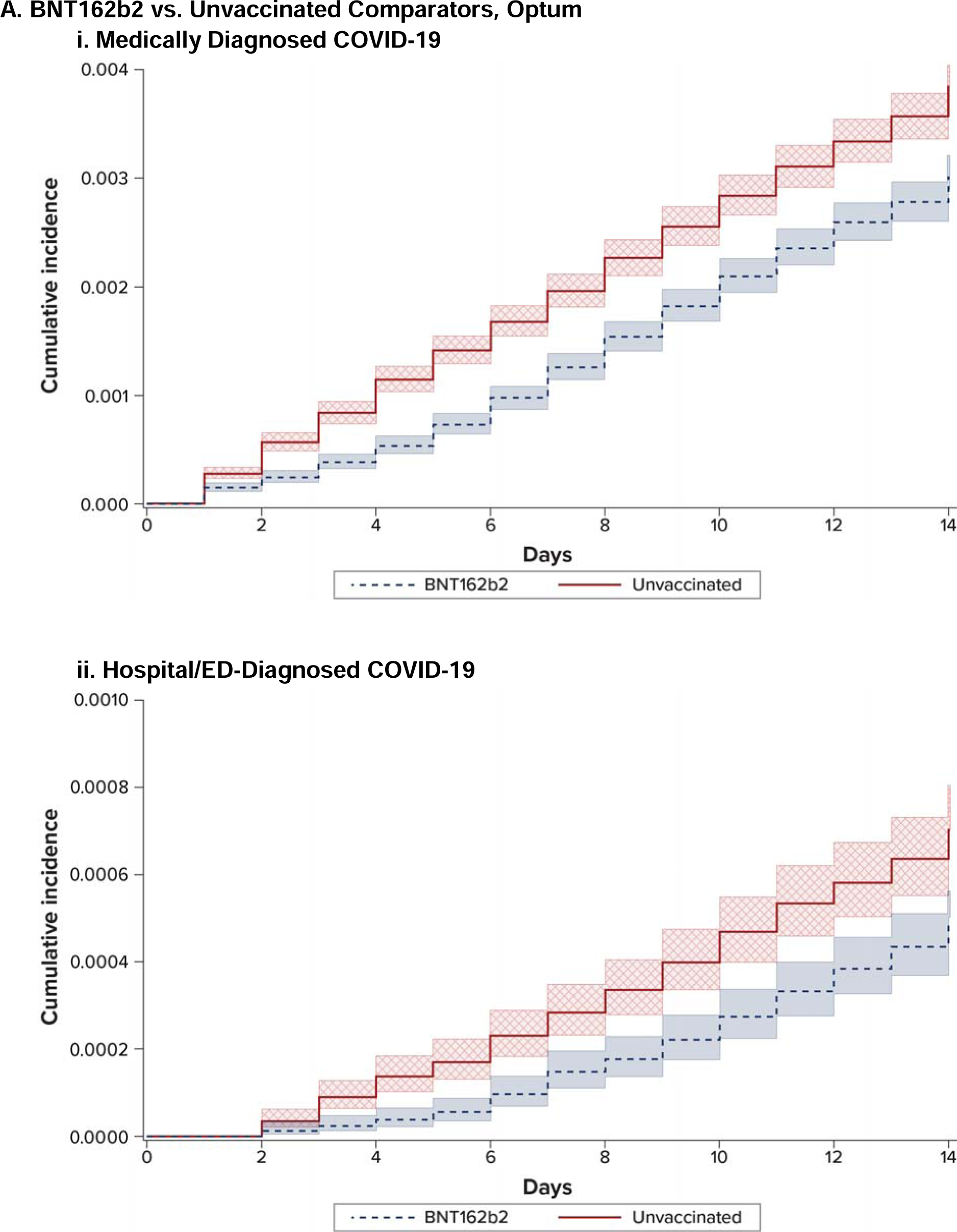
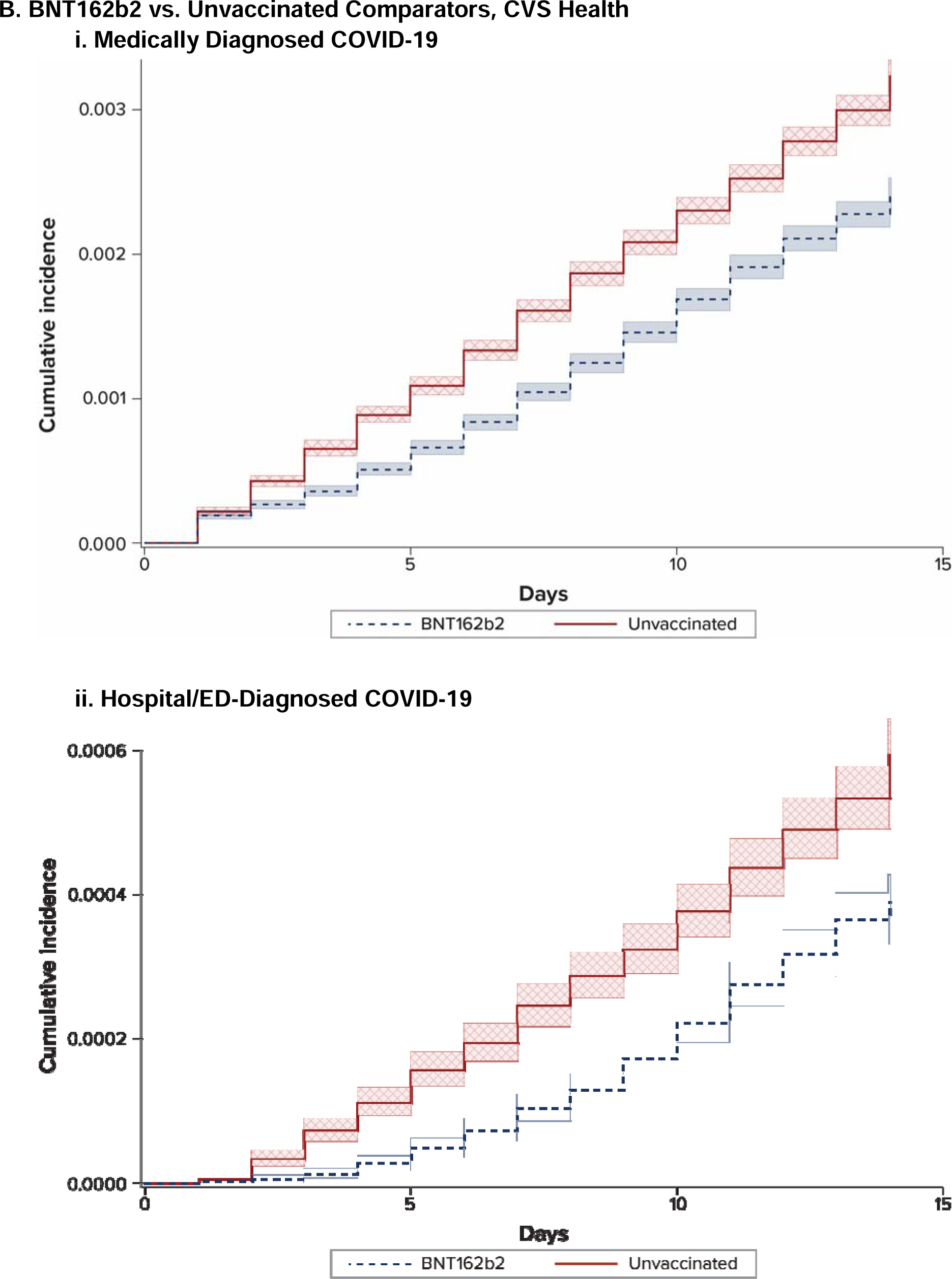
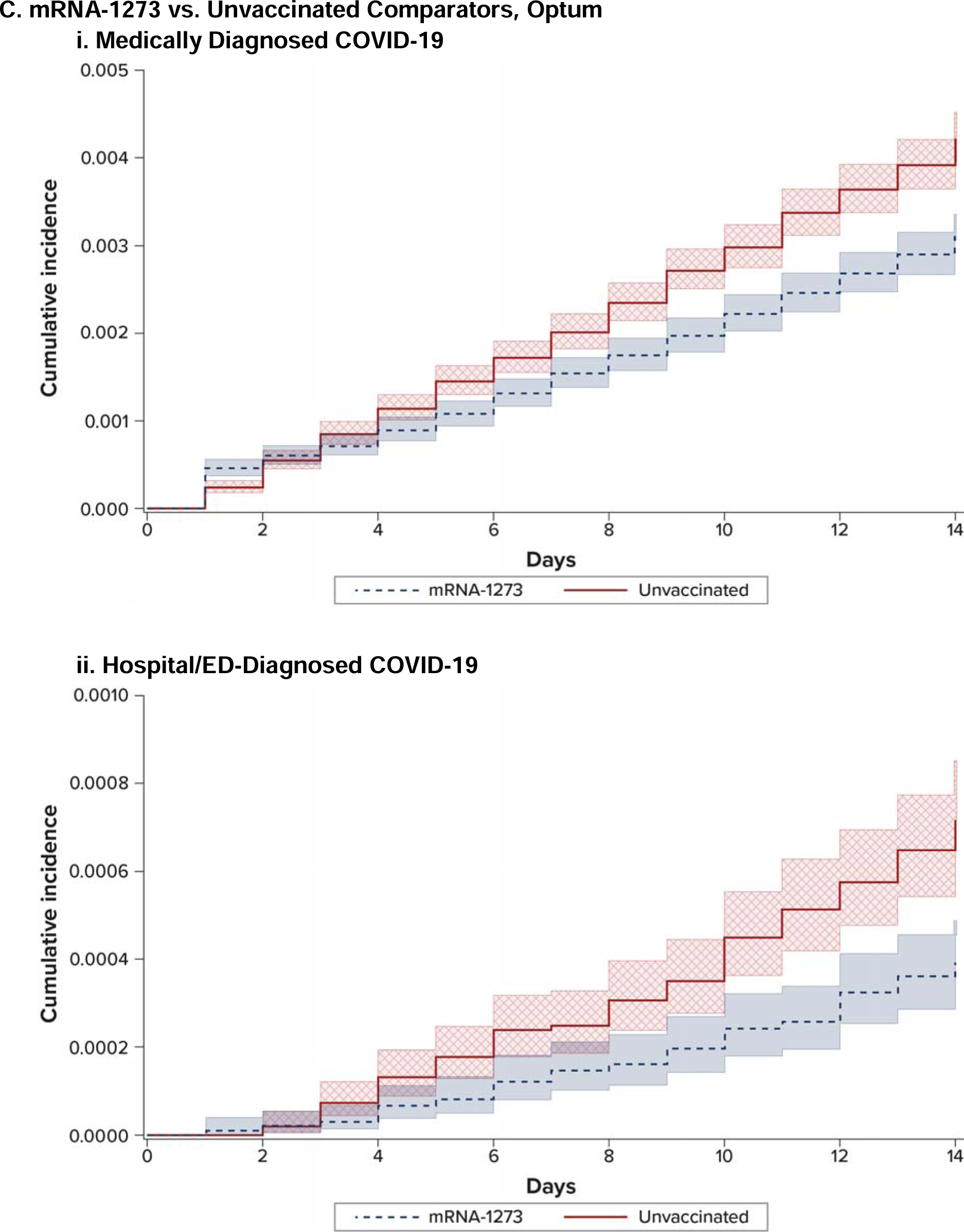
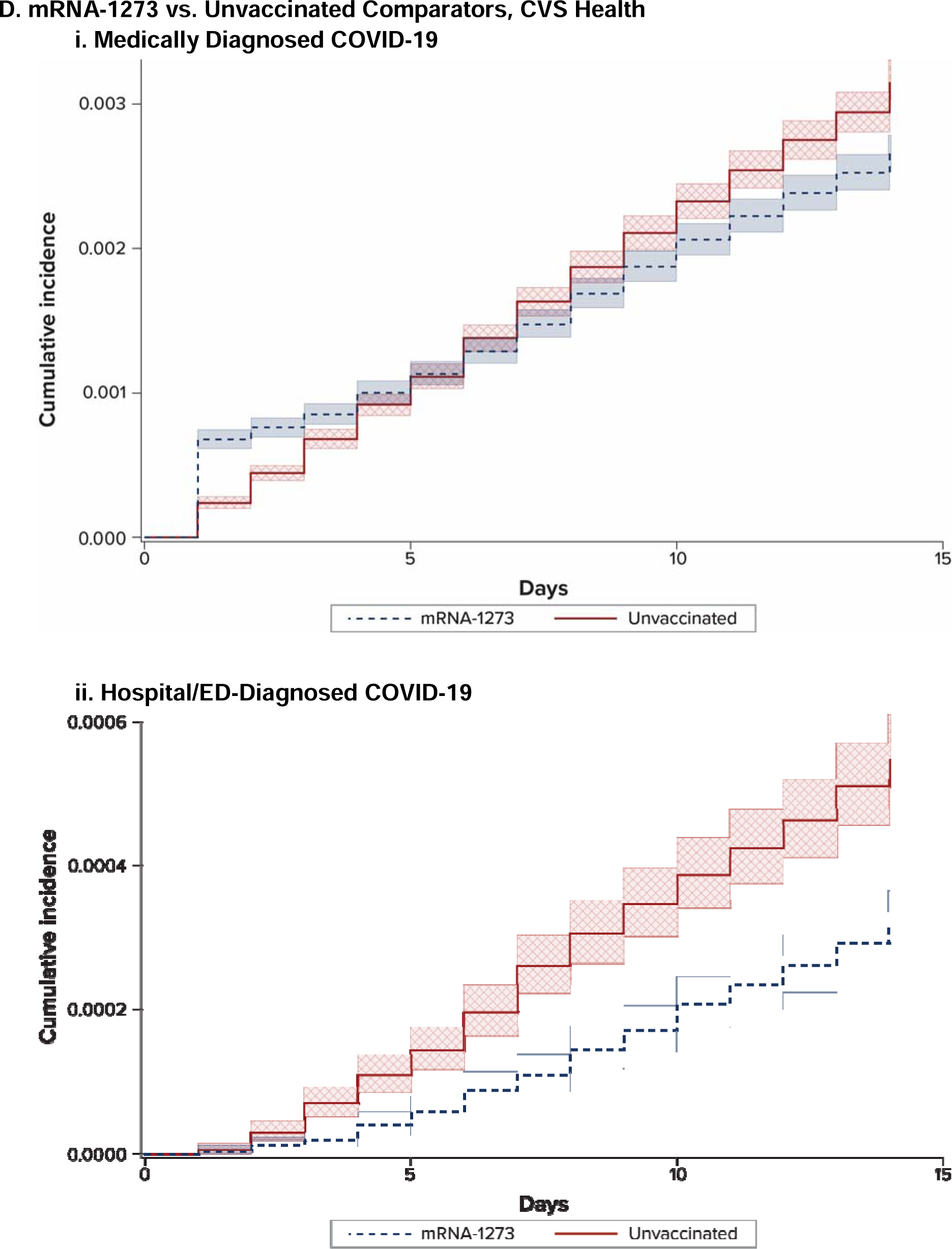
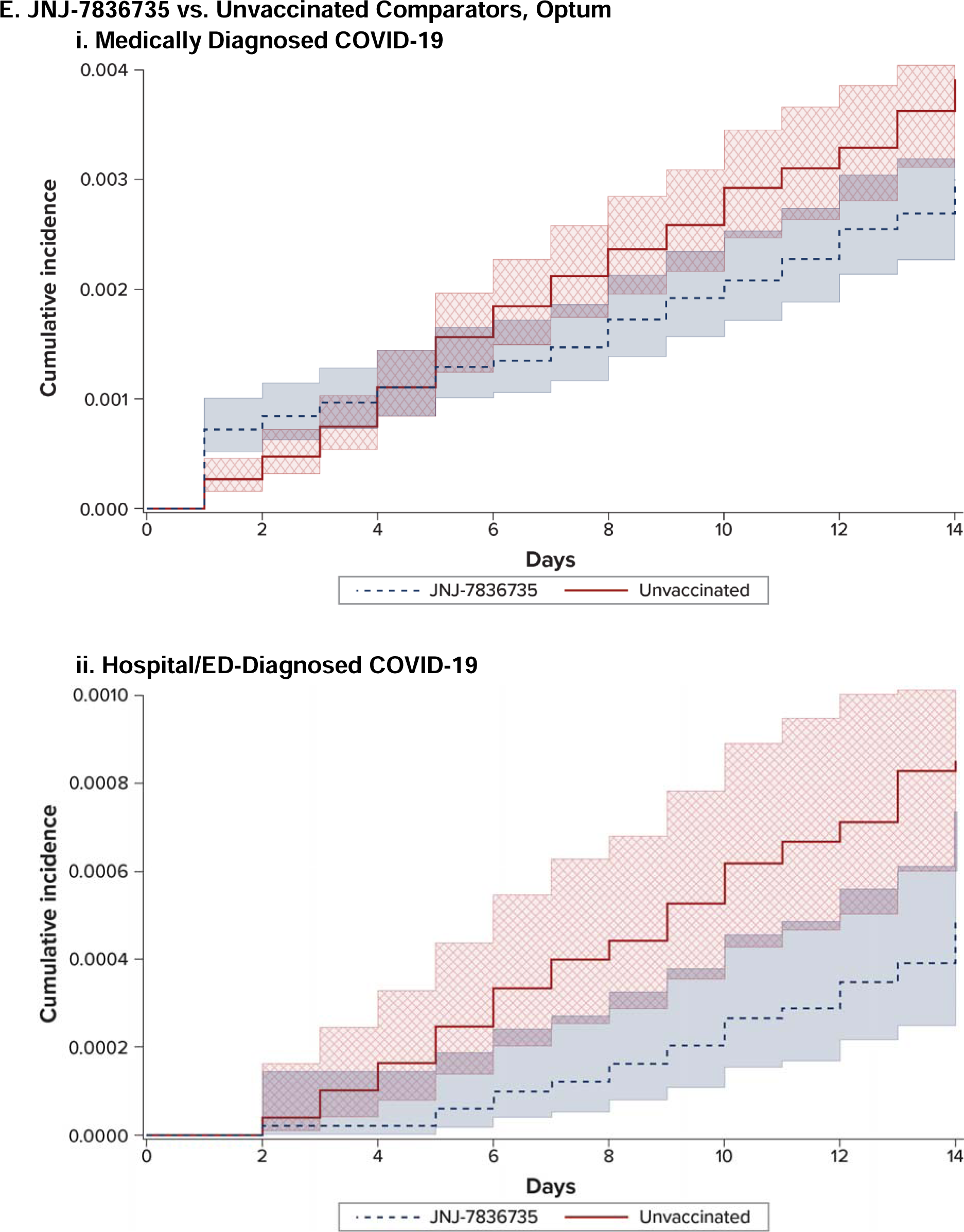
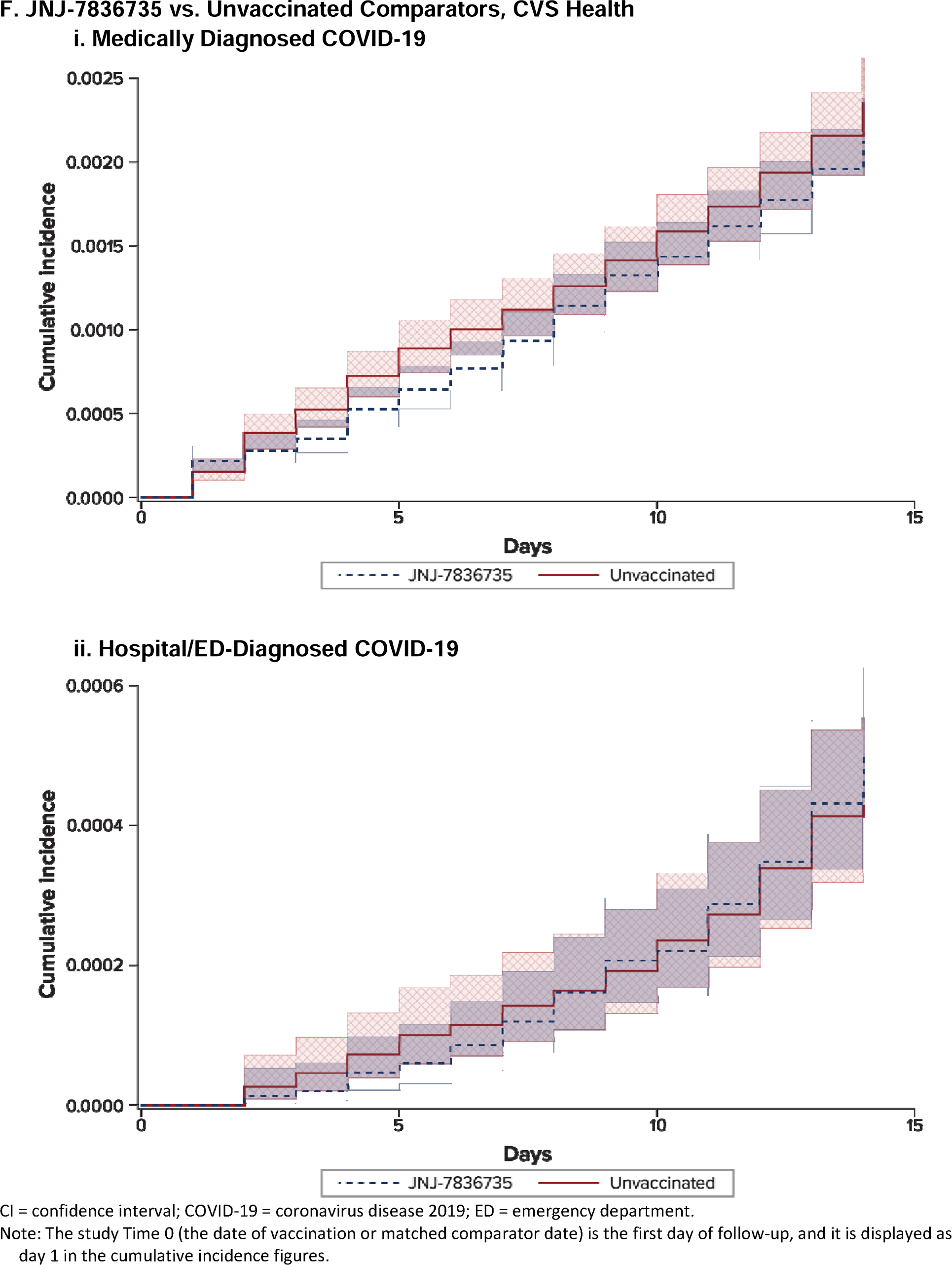
Cumulative Incidence of COVID-19 Outcomes in Adults Receiving a Complete Primary Series of COVID-19 Vaccines and Matched Unvaccinated Comparators, Negative Control Outcome in the First 14 Days.

**sFigure 6.**
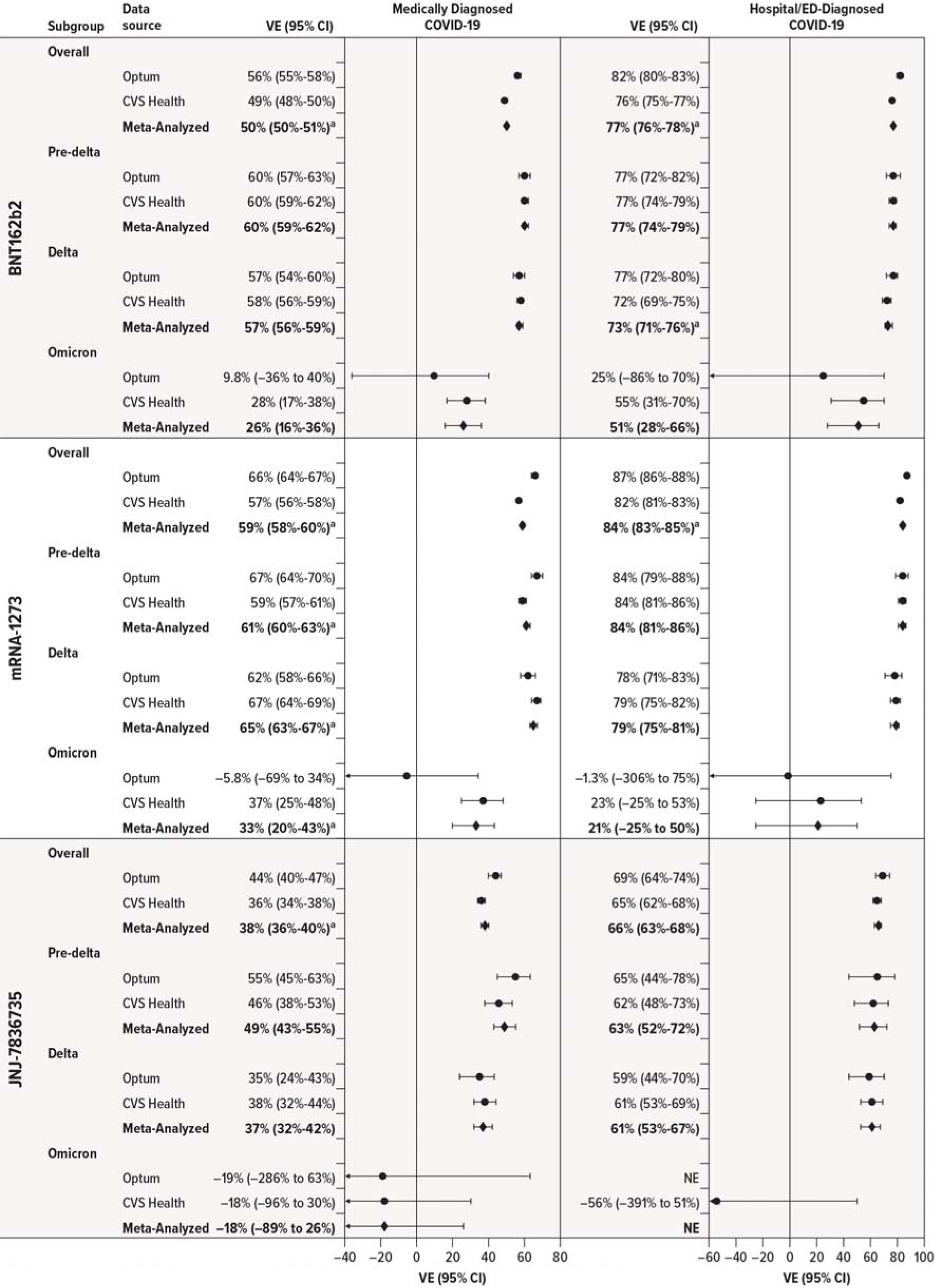
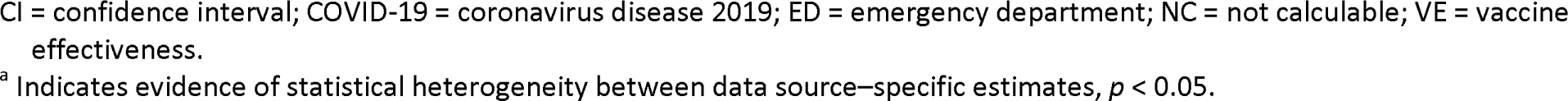
Estimated Effectiveness of Receiving a Complete Primary Series of COVID-19 Vaccine in Adults Aged 18-64 Years, Compared With Being Unvaccinated, by SARS-CoV-2 Variant Era.

**sFigure 7.**
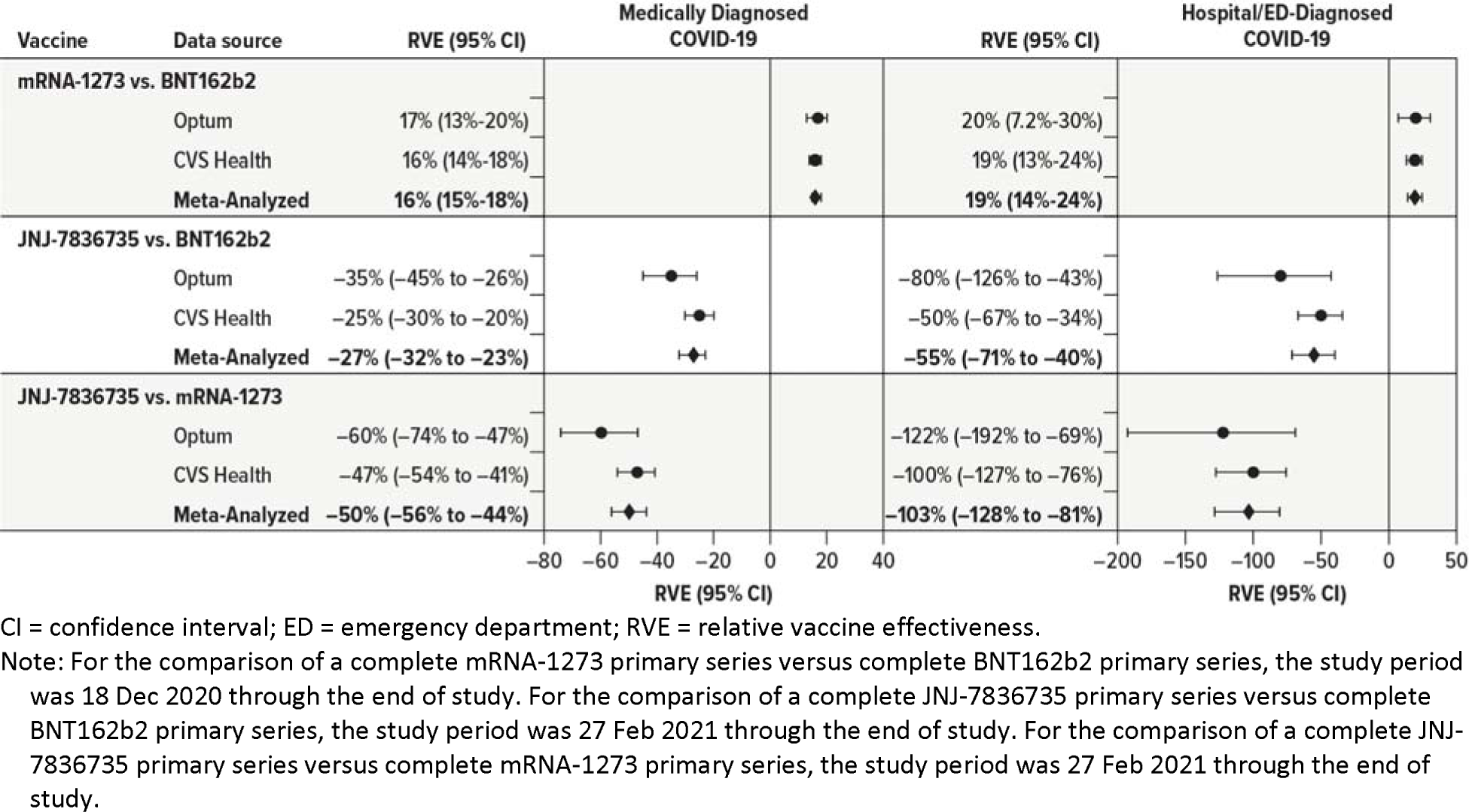
Estimated Relative Effectiveness of Receiving a Complete Primary Series of COVID-19 Vaccine in Adults Aged 18-64 Years, Compared With Receiving a Complete Primary Series of Different COVID-19 Vaccines.

**sFigure 8.**
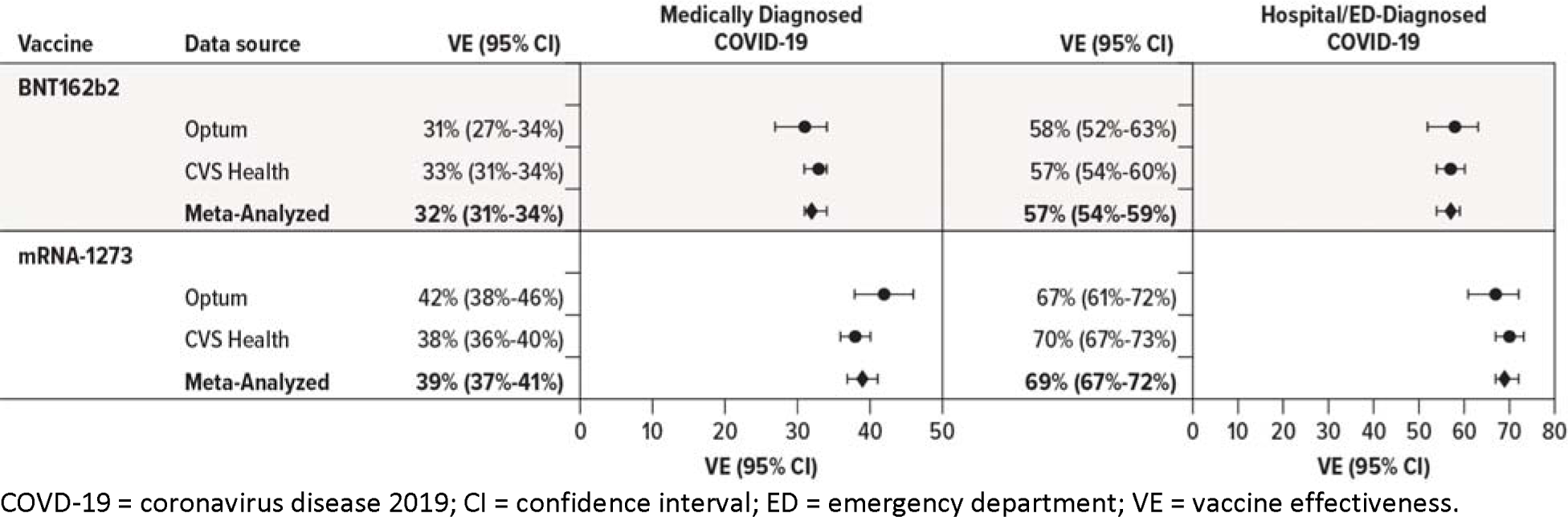
Estimated Effectiveness of Receiving a Single Dose of a 2-Dose Primary Series of COVID-19 Vaccine in Adults Aged 18-64 Years, Compared With Being Unvaccinated.

## Notes

### Competing Interest Statement

This work was supported by the United States Food and Drug Administration as part of the BEST Initiative. JBL, CB, DI, MMR, and MSA are employees of RTI International, an independent, nonprofit research institute that performs research on behalf of governmental and commercial clients, including vaccine manufacturers. LP, JD, RO, RP, MM, JS, LBW, GWY, JS, and KA are employees of Optum and own stock in UnitedHealth Group. EJB is an employee of Optum. DAD and CNMW are employees of CVS Health. The other authors have no conflicts to report.

### Clinical Protocols

https://bestinitiative.org/wp-content/uploads/2022/03/C19-VX-Effectiveness-Protocol_2022_508.pdf

### Author Declarations

This activity was conducted as part of the FDA public health surveillance mandate, and FDA did not require Institutional Review Board review.

